# Identification of neuropathology-based subgroups in multiple sclerosis using a data-driven approach

**DOI:** 10.1101/2023.05.15.23289980

**Authors:** Alyse de Boer, Aletta M.R. van den Bosch, Nienke J. Mekkes, Nina Fransen, Eric Hoekstra, Joost Smolders, Jörg Hamann, Inge Huitinga, Inge R. Holtman

## Abstract

Multiple sclerosis (MS) is a heterogeneous disorder with regards to clinical presentation and pathophysiology. Stratification into biologically distinct subgroups could enhance prognostication and efficacious allocation to disease-modifying therapies. In this study, we identified MS subgroups by performing a clustering analysis on neuropathology data collected for MS donors in the Netherlands Brain Bank (NBB) autopsy cohort. The input dataset contained detailed information on white matter lesion load, the proportion of active, mixed active/inactive, inactive and remyelinating lesions, microglia morphology in these lesions, and the presence of microglial nodules, perivascular cuffs and cortical lesions for 228 donors. A factor analysis was performed to reduce noise and redundancy prior to hierarchical clustering with K-means consolidation. Four subgroups with distinct patterns of white matter lesions were identified. These were subsequently validated with additional clinical, neuropathological and genetic data. The subgroups differed with regards to disease progression and duration, the timing of motor, sensory and other relevant signs and symptoms, patterns of cortical lesions and the presence of B cells. Age at MS onset and sex, previously associated with milder forms of MS, did not differ between the subgroups; the subgroups could also not be distinguished based on the manifestation of clinical signs and symptoms. The available genetic data was used to calculate MS polygenic risk scores (PRSs) for donors included in the NBB cohort. The MS PRS did not differ between the subgroups, but was significantly correlated with the first and second dimension of the factor analysis, the latter lending genetic support to our subdivision. Taken together, these findings suggest a complex relationship between neuropathological subgroups and clinical characteristics, indicating that post-mortem cohort studies are critical to better stratify patients and understand underlying neuropathophysiological mechanisms, in order to ultimately achieve personalised medicine in MS.

## Introduction

Multiple sclerosis (MS) is one of the most common neurological disorders in early adulthood, resulting from a combination of environmental, lifestyle and genetic factors [53, 55]. This chronic disease is characterized by lesions in the central nervous system, with varying extents of inflammation, demyelination, loss of axons and gliosis [28]. Since many different regions in the brain and spinal cord can be affected, MS is associated with a broad range of clinical signs and symptoms related to neurological dysfunction. In addition, there is marked variation in disease course, severity and response to disease-modifying treatment [26, 45]. Historically, MS has been divided into three clinical phenotypes: relapsing remitting (RR), primary progressive (PP) and secondary progressive (SP) MS [32, 33]. This subdivision neither considers the possibility of relapses in progressive MS, nor progression independent of relapse activity in relapsing MS [6]. Therefore, the currently leading view is that this clinical division is rather artificial and that MS should be seen as one heterogeneous disease entity, best described in terms of relapse activity and progression [29, 32].

To address the heterogeneity of MS and the challenge this represents for allocating treatment and providing an accurate prognosis, multiple studies have defined MS subtypes based on underlying pathobiological mechanisms, using theory-driven as well as data-driven approaches. For example, an analysis of lesion-level magnetic resonance imaging (MRI) data led to the identification of two patient clusters, which were associated with different levels of clinical disability [50]. Another MRI study defined three subtypes, termed cortex-led, normal-appearing white matter-led and lesion-led after the site of the earliest MRI abnormalities, which differed in the progression of disease and response to treatment [13]. Furthermore, a study investigating neuropsychological data found five distinct cognitive phenotypes, which were related to clinical, demographic and MRI features and could be of added value in clinical care [12]. With regards to immunopathology, four patterns of demyelination in early, active MS lesions have been reported [34]. The patterns were heterogeneous between individuals, while lesions within the same brain generally followed the same patterns, even over time [34, 42]. This suggests that the four patterns could be used to define MS subgroups that correspond with distinct pathogenetic mechanisms involved in demyelination [34, 42]. However, it should be noted that this particular subdivision does not hold in more advanced MS as studied at autopsy [5], which is the topic of this study.

Here, we aimed to identify data-driven neuropathology-based subgroups, by applying a clustering analysis to post-mortem neuropathology data collected for MS brain donors in the Netherlands Brain Bank (NBB) autopsy cohort. The in-depth characterization of MS brain tissue facilitated a unique input dataset, containing information on the number of white matter lesions and reactive sites, the proportion of active, mixed active/inactive (mixed), inactive and remyelinated lesions, the morphology of microglia present in active and mixed lesions, and the presence of microglial nodules, perivascular cuffs and cortical lesions. To investigate the validity of the resulting subgroups, additional neuropathological, clinical and genetic data available for this cohort were assessed. Key parameters that we included were related to disease progression and duration, signs and symptoms and their temporal pattern, gray matter lesions, the presence of B cells in the brainstem, MS-associated genetic variants and polygenic risk scores (PRSs).

## Methods

### NBB autopsy procedures and MS lesion characterization

NBB donors provided informed consent for brain autopsy and the use of tissue and data for research purposes. The procedures of the NBB are in compliance with Dutch and European law and have been approved by the Ethical Committee of the VU University Medical Center in Amsterdam, the Netherlands.

Both standardized locations in the brainstem and spinal cord as well as macroscopically detectable MS lesions were dissected, as previously described by Luchetti et al. [35]. Since 2001, lesions were additionally dissected under guidance of post-mortem MRI, which increased the yield of active demyelinating lesions and reactive sites [11]. Lesions were classified based on double immunostaining for proteolipid protein and human leukocyte antigen (HLA-DR-DQ) [35]. In total, five qualitatively different white matter lesion types were distinguished: I) reactive sites, clusters of HLA^+^ microglia/macrophages without signs of demyelination; II) active lesions, with partial demyelination and accumulation of HLA^+^ cells throughout the lesions; III) mixed lesions, with a demyelinated, hypocellular and gliotic centre, and a rim of HLA^+^ cells; IV) inactive lesions, a fully demyelinated, hypocellular and gliotic region with no accumulation of HLA^+^ cells; and V) remyelinated lesions, with partial myelination and few HLA^+^ cells. The HLA^+^ microglia/macrophages in active and mixed lesions were additionally scored for morphology: I) thin and ramified; II) amoeboid (rounded) with few ramifications and III) foamy. Gray matter lesions were dissected as well, and classified according to their location into: I) leukocortical, located in both white and gray matter; II) intracortical and III) subpial. Lesions were only considered subpial if the tissue block contained the first layer of the cortex, with the consequence that the number of intracortical lesions is overestimated to some extent.

Lesion load was previously defined as the total number of active, mixed, inactive and remyelinated lesions present in standardly dissected brainstem tissue blocks; similarly, reactive site load was calculated as the total number of reactive sites in these brainstem blocks [35]. These values are therefore not affected by sampling bias and considered to be an adequate reflection of the total number of white matter lesions present in a donor. Furthermore, the proportions of active, mixed, inactive and remyelinated lesions relative to the total number of white matter lesions in all dissected tissue blocks were calculated [35]. In addition, the tissue blocks were assessed to determine the presence of cuffs (defined as >1 perivascular ring of leukocytes) and microglia nodules. This was converted into binary scores per donor, indicating the overall presence or absence of cuffing and nodules, respectively.

Cortical lesions were scored as either present or absent, and considered present when at least one cortical lesion was identified in the cortical blocks dissected of that donor. If one or more cortical lesions were present, the sum and relative proportions of the different gray matter lesion types were determined. To quantify the cortical lesion load of a donor, the number of lesions was divided by the total number of cortical tissue blocks investigated for that donor. Note that in contrast to the white matter lesion load, this value can be affected by sampling bias.

### Identification of neuropathology-based subgroups

#### Pre-processing of neuropathology data

A centered log ratio (CLR) transformation was applied to the proportional lesion data, to reduce the possibility of correlations arising solely from the constant sum constraint (i.e. for each donor, the values of the proportions sum to 1) [20]. The CLR transformation takes the logarithm of each lesion proportion, relative to the geometric mean of all lesion proportions for a donor; because this requires non-zero data, multiplicative zero replacement was performed beforehand [38]. Both were implemented using the package Compositional (v5.6) with R version 4.1.3 [49].

Analysis was restricted to donors for whom complete proportional white matter lesion data was available (n = 228). To maximize the number of observations in the dataset, missing data in other variables was accepted. For 14 donors, lesion load and reactive site load were missing, most probably because no standard locations were dissected. Data on the presence of cortical lesions was missing in 20 cases. For these donors, it was not known whether no cortical lesions were identified because they were not present or because no blocks with cortical tissue had been dissected. All missing values were imputed with the missMDA package (v1.18) [25], based on 12 dimensions (explaining all variance). The imputed dataset consisted of 228 observations for 13 neuropathology-related variables: transformed white matter lesion proportions, lesion load and reactive site load and the presence or absence of cuffing, nodules and cortical lesions. The final pre-processing step consisted of a factor analysis of mixed data (FAMD) using the FactoMineR package (v2.4) [31], which scales continuous and categorical variables in order to balance their impact on the analysis.

#### Clustering analysis

Agglomerative hierarchical clustering was performed on the subset of dimensions with an eigenvalue > 1, in accordance with the Kaiser-Guttman criterion. In this way, FAMD served to denoise the data [23]. The optimal number of clusters was determined from the hierarchical tree; partitional clustering with the K-means algorithm was then performed to further consolidate the clusters [23]. Both types of clustering were performed with FactoMineR (v2.4).

### Validation with additional neuropathology, clinical and genetic data

To interpret the meaning and validity of the clusters, relevant external data, not used as input for the clustering algorithm, is of key importance [1]. Here, we assessed whether the clusters displayed differences in neuropathological, clinical and genetic data.

#### Description of general clinical data

General clinical information was obtained by retrospective chart analysis for the majority of the donors [35]. Clinical course was extracted from the patient files, categorised by the treating neurologist in clinical practice into relapsing without disease progression (RR), progressive with a relapsing onset (SP) or progressive without dominant relapsing onset (PP). These phenotypes were reviewed by an independent neurologist. Other characteristics determined in this manner were age at onset, time from onset to Expanded Disability Status Scale (EDSS)-6 and duration of disease.

#### Analysis of signs and symptoms and their temporal patterns

For a complete description of the processing of clinical text data and subsequent visualisation, see Mekkes et al. [41]. Briefly, state-of-the-art natural language processing techniques were used in order to identify 80 signs and symptoms in individual sentences, which led to a high-quality dataset of clinical disease trajectories for 3081 NBB donors, showing clear disease specific profiles. A clinical disease trajectory was available for 219 of the 228 donors in this study. This allowed us to determine the mean number of observations per sign/symptom and the proportion of donors with at least one observation for the MS subgroups as well as a group of control NBB donors without neurological diseases (n = 444; referred to as non-diseased controls). A permutation test was performed to assess which signs and symptoms were significantly overrepresented compared to a background of NBB donors with clearly defined neuropathological diagnoses (n = 2061; termed non-MS controls). Moreover, the temporal distribution of observations of a subset of signs and symptoms relevant to MS was determined for the MS subgroups and non-diseased controls.

#### Description of neuropathological data

Whereas the binary variable related to the presence/absence of cortical lesions was part of the input dataset, the proportions of gray matter lesion types were used for validation. Similar to the white matter lesion proportions, a CLR transformation was applied before comparing the different subgroups. Furthermore, information on the presence of B cells in the medulla oblongata was available for 132/228 donors, from a previous study by Fransen et al [17].

#### Genotyping of the NBB autopsy cohort, quality control (QC) and imputation

Samples derived from donors in the NBB autopsy cohort were genotyped with the Infinium Global Screening Array (Illumina, v3) by the Human Genomics Facility at Erasmus Medical Centre. Unless otherwise specified, all processing steps and analyses were performed with the PLINK toolset version 1.90 beta 6.24 [48]. Pre-imputation QC consisted of iterative removal of variants and samples with missing data so that the final call rate exceeded 99%, exclusion of samples with excess heterozygosity (inbreeding coefficient F_sample_ < mean F − 4 × standard deviation (SD)) and removal of variants deviating from Hardy-Weinberg equilibrium (P < 1 × 10^−5^). To improve detection of rare variants, zCall was applied in a post-processing step followed by QC using the same thresholds [19]. Imputation was performed in a two-step procedure using SHAPEIT for phasing and Minimac4 for imputation to the HRC (Haplotype Reference Consortium) r1.1 reference panel [9, 40]. Monomorphic and ultra-rare variants (minor allele frequency < 0.005) were removed. Principal component analysis on ancestry informative markers of samples included in this study and the 1000 Genomes Project (phase 3, v5) identified 44 NBB donors with non-European ancestry (deviating > 4 × SD from the mean of the European reference dataset on the first four principal components), which were excluded from further analysis [52]. Correction for family relations within the NBB cohort was performed by removing one individual per pair of samples with genomic relatedness > 0.025.

#### Genetic analyses

For 192/228 MS donors genotype data was available. Other brain donors in the NBB autopsy cohort served as controls, except those with possible (but not definite) MS or a diagnosis of neuromyelitis optica (NMO). The latter were excluded because MS risk variants have been associated with NMO susceptibility [39]. Of the 2049 non-MS non-NMO controls, 257 (12.5%) were non-demented. The majority had been diagnosed with one or more neurological disorders, most commonly Alzheimer’s disease (n = 718, 35.0%), Parkinson’s disease (n = 233, 11.4%) and/or frontotemporal dementia (n = 179, 8.7%); 119 (5.8%) donors had a psychiatric disorder as main diagnosis. Since the genetic correlation of these brain disorders with MS is limited, this does not preclude their suitability as control [2]. For a further description of the controls, see supplemental Table 1 (Online Resource 1). The most recent MS genome-wide association study (GWAS) by Patsopoulos et al. [47] identified 200 independent, autosomal variants outside the major histocompatibility complex (MHC) region that were the best marker for the true underlying effects on MS susceptibility. Genotype information was directly available for 183/200 of these variants. Using the LDlink tool (release 5.5.1; European reference population), 9 single nucleotide polymorphisms (SNPs) in strong linkage disequilibrium (LD; defined as r^2^ > 0.8) with one of the remaining 17/200 variants were identified and included as well [36]. With regards to the MHC region, the tagging SNP for the HLA-DRB1*15:01 allele was included in the analysis (rs3135388) [10, 21]. Furthermore, the SNPs previously identified by Fransen et al. [16], related to MS severity and pathology, were selected. To ensure that the SNPs in the final selection were biallelic and not in high LD with each other, 8 variants were excluded with the LDlink tool (see suppl. Table 2, Online Resource 1, for a detailed overview of the 288 SNPs in the final selection). A standard association analysis (chi-square allelic test) was performed with PLINK, comparing each neuropathology-based subgroup with controls for the 288 MS-associated variants. In addition, MS PRSs were constructed using the LDAK-BayesR-SS tool described by Zhang et al. [58]. We implemented QuickPRS to construct a prediction model based on the GWAS by Patsopoulos et al. [47].

### Statistical analysis

All statistical analyses were performed in R (v4.1.3) or Python (v3.8.8). Kruskal-Wallis and Mann-Whitney tests or Fisher’s Exact test were used for neuropathological, clinical and genetic (PRS) data. Bonferroni correction for multiple comparisons between the subgroups was performed per variable (unless otherwise specified). With regards to the distribution (dot plot) and temporal pattern (violin plot) of clinical signs and symptoms, the Benjamini Hochberg - False Discovery Rate was applied, with the significance threshold set to p < 0.1 and p ≤ 0.01, respectively, as previously described [41]. Correlation between PRS and neuropathological dimensions was assessed by performing linear least-squares regression and calculating Pearson’s correlation coefficient.

## Results

### MS donors can be divided into four subgroups with distinct patterns of white matter lesions

To identify subgroups of patients according to shared neuropathological features, we established a computational workflow, shown in Fig. 1a. A factor analysis of mixed data was performed and the first five dimensions, together explaining 62% of the total variance (suppl. Table 3, Online Resource 2), were selected as input for clustering. The first dimension reflects a spectrum from low to high demyelinating activity, ranging from a high lesion load with active and mixed lesions populated by foamy microglia on one hand, and a predominance of inactive and remyelinated lesions with few HLA^+^ microglia/macrophages on the other hand (Fig. 1b). The second dimension is positively related to the number of reactive sites, presence of nodules and proportion of active lesions with ramified microglia, and inversely to mixed lesions with foamy microglia. The third dimension correlates with a higher proportion of mixed lesions with low microglia activity scores and a lower proportion of active lesions with foamy microglia. Variables related to active and mixed lesions with rounded microglia contribute mainly to dimension 4, while dimension 5 mostly reflects the presence of perivascular cuffing.

**Fig. 1.**
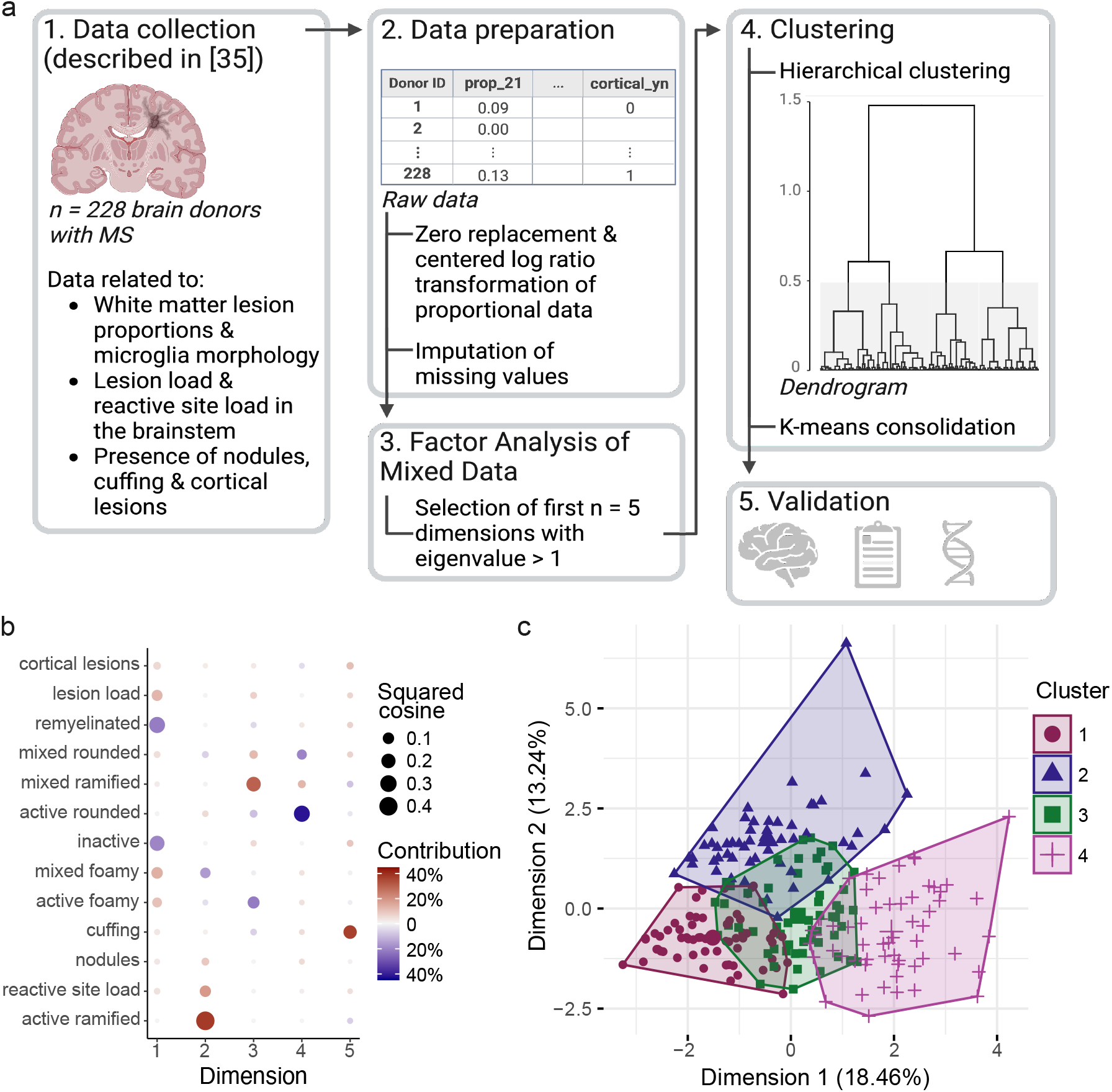
MS donors can be divided into four subgroups based on white matter neuropathology. **a** Schematic overview of the computational workflow, created with BioRender.com. **b** Dot plot displaying the relation between the input variables for the factor analysis and the first five dimensions. Dot size indicates the squared cosine, reflecting the proportion of variance in a variable explained by a dimension. Higher values correspond to a better quality of representation of the variable by the dimension. Dot color indicates whether the correlation between the dimension and the variable is positive (red) or negative (blue); color intensity reflects the relative contribution of the variable to the component. **c** Factor map with the MS donors (n = 228) plotted on dimension 1 and 2. Color and point shape indicate the cluster a donor belongs to. The explained variance of the dimension is given between brackets in the axis labels. For factor maps of the MS donors on the remaining dimensions, see suppl. Fig. 1

The subsequent hierarchical clustering step clearly revealed four different MS subgroups (dendrogram in Fig. 1a), which were consolidated using K-means clustering. Cluster 1 (n = 63) and 4 (n = 57) are well separated on the first dimension (Fig. 1c); donors in cluster 2 (n = 52) and 3 (n = 56) score high on the second and third dimension, respectively (Fig. 1c and suppl. Fig. 1, Online Resource 3). For a complete overview of how the input variables differ between the subgroups, see suppl. Fig. 2 and 3 (Online Resource 3). Briefly, cluster 1 is characterized by a higher proportion of inactive and remyelinated lesions compared to the other clusters, in combination with a lower reactive site load and lesion load in the brainstem. Moreover, the majority of MS donors without cortical lesions are part of this subgroup. Cluster 2 has a higher proportion of active lesions with ramified and rounded microglia and relatively more remyelinated lesions than cluster 3 and 4. Compared to cluster 1 and 2, the ratio of remyelinated to inactive lesions is low for donors in cluster 3. Cluster 3 is further characterized by a relatively high number of mixed lesions with ramified and rounded microglia. MS donors in cluster 4 have the highest proportion of lesions with foamy microglia (both active and mixed) and the lowest proportion of inactive lesions relative to donors in the other clusters. The four MS subgroups were validated using orthogonal data types, as described in the remainder of this section.

**Fig. 2.**
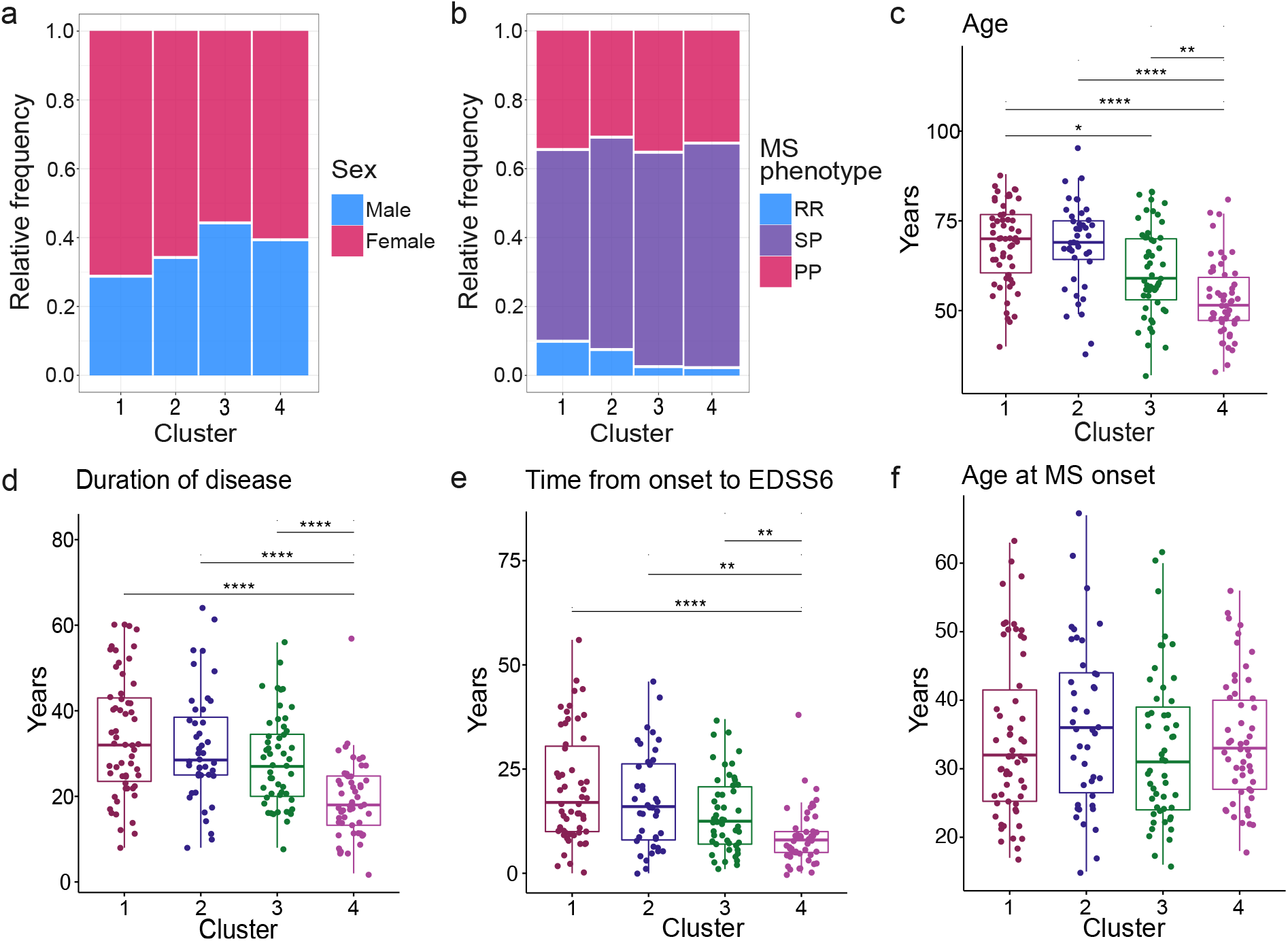
Demographic and clinical variables per MS subgroup. **a** Mosaic plot showing the proportion of males and females. Sex was known for 215 donors (1: n = 63; 2: n = 44; 3: n = 52; 4: n = 56). There are no significant differences (Fisher’s exact test; p = 0.34). **b** Mosaic plot showing the proportion of donors with RR, SP and PP MS. MS phenotype was determined for 197 donors (cluster 1: n = 52; 2: n = 42, 3: n = 48; 4: n = 55). Clusters do not differ significantly (Fisher’s Exact test; p = 0.59). **c** Box plot of age at death for 205 donors (1: n = 58; 2: n = 42; 3: n = 51; 4: n = 54). **d** Box plot of duration of disease for 207 donors (1: n = 58; 2: n = 44; 3: n = 51; 4: n = 54). **e** Box plot of years from MS onset to EDSS-6 for 194 donors (1: n = 55; 2: n = 40; 3: n = 50; 4: n = 49). **f** Box plot of age at MS onset for 204 donors (1: n = 58; 2: n = 42; 3: n = 51; 4: n = 53). Significance in **c-f** was assessed by pairwise Mann-Whitney tests and adjusted for multiple testing (Bonferroni). *p ≤ 0.05; **p < 0.01; ***p < 0.001; ****p < 0.0001

**Fig. 3.**
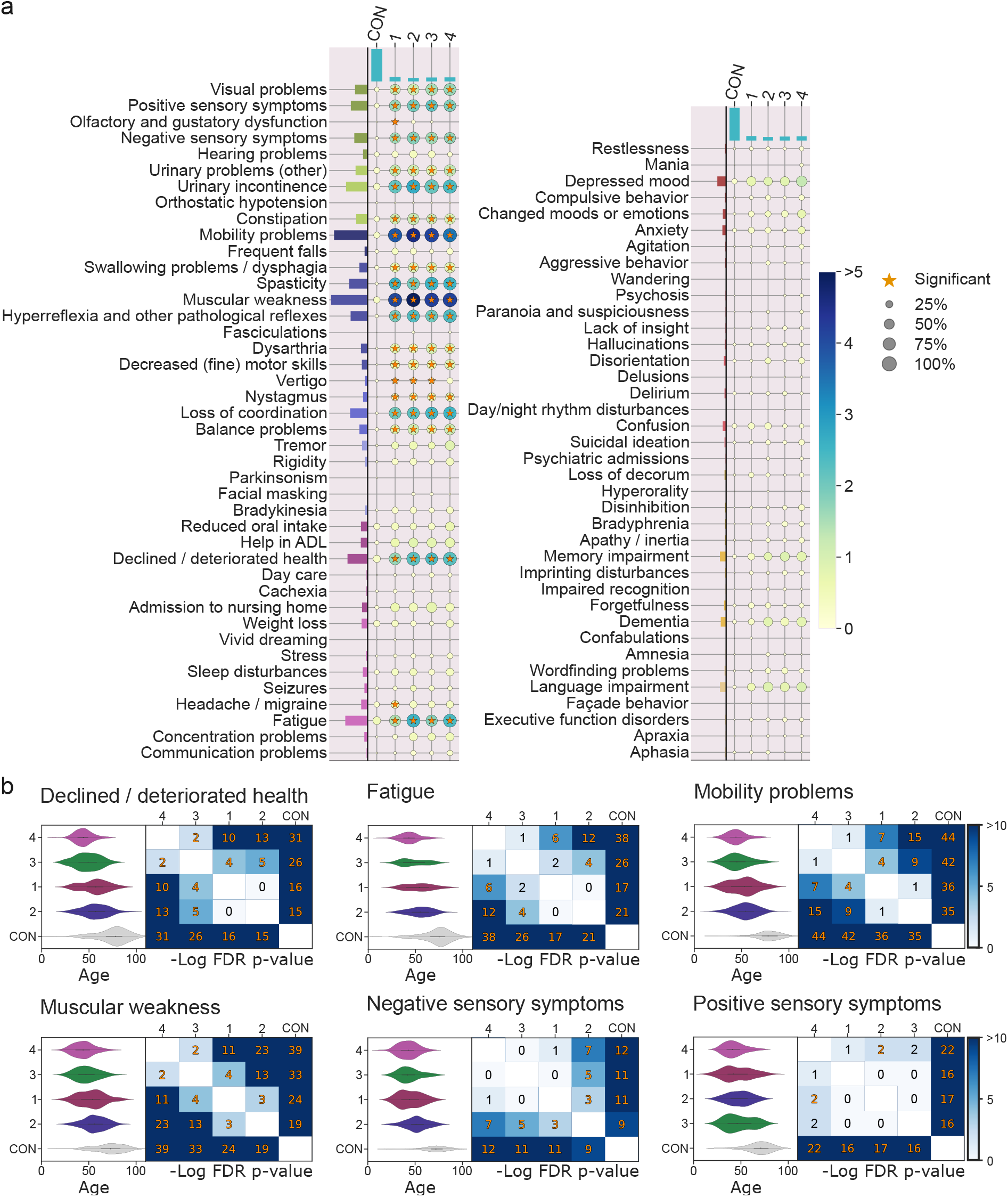
Manifestation, frequency and temporal pattern of clinical signs and symptoms. **a** Integrated dot and bar plot showing the manifestation of 80 signs and symptoms (Y-axis) for non-diseased NBB controls and MS subgroups 1-4 (X-axis). The dot size corresponds to the proportion of donors in which a sign/symptoms was observed. The dot color corresponds to the mean number of observations of a sign/symptom across donors. Significantly overrepresented signs/symptoms were visualized with an asterisk (FDR corrected p < 0.1). **b** Violin plots showing the temporal distribution of motor, sensory and other symptoms for the MS subgroups and non-diseased NBB controls. The heatmap shows the results of pairwise Mann-Whitney tests, with significant -10log FDR corrected p-values in orange (p ≤ 0.01). ADL = activities of daily living; FDR = false discovery rate; CON = (non-diseased) controls

### MS donors in cluster 4 have a severe disease course

To better understand the clinical implications of the identified subgroups we subsequently focused our analysis on data types related to the autopsy procedure, demographics and disease experience. Of note, the subgroups do not differ with regards to post-mortem delay (p = 0.20), pH (p = 0.69) or brain weight (p = 0.95) (suppl. Table 4, Online Resource 2), suggesting that the clusters are not driven by these covariates. Interestingly, there is also no significant difference in sex (p = 0.34) or MS clinical phenotype (p = 0.59) between the subgroups (Fig. 2a,b), the latter supporting the notion that the historically identified clinical MS phenotypes do not qualitatively differ with regards to pathology.

Year of death does differ between the subgroups (p = 6.7 × 10^−4^). The autopsy of donors in cluster 2 was generally performed more recently than of donors in cluster 1 (p = 1.7 × 10^−4^) and cluster 4 (p = 0.044) (suppl. Fig. 4a, Online Resource 3). This means that donors in cluster 2 more often had an MRI-guided autopsy and probably received more recent forms of therapy, which may have affected the number and type of lesions identified and thereby the clustering results. In addition, the cause of death varies between the subgroups (Fisher’s Exact, simulated p ≈ 0.012; suppl. Fig. 4b, Online Resource 3). Although donors in all four subgroups mostly died of natural causes, legal euthanasia was relatively more common in cluster 2 and 4 compared to the other subgroups. Furthermore, patients in cluster 4 died at an earlier age (1 vs 4: p = 4.3 × 10^−9^; 2 vs 4: p = 2.6 × 10^−7^; 3 vs 4: p = 0.003; Fig. 2c), after a shorter disease duration (1 vs 4: p = 4.9 × 10^−8^; 2 vs 4: p = 1.6 × 10^−6^; 3 vs 4: p = 6.9 × 10^−5^; Fig. 2d). The time between MS onset and reaching EDSS-6 is shorter for this subgroup (1 vs 4: p = 1.5 × 10^−6^; 2 vs 4: p = 0.001; 3 vs 4: p = 0.006; Fig. 2e), indicating a higher disease severity. Median age at MS onset ranges from 31 years for donors in cluster 3 to 36 years for cluster 2, with no significant differences between the subgroups (p = 0.47; Fig. 2f).

**Fig. 4.**
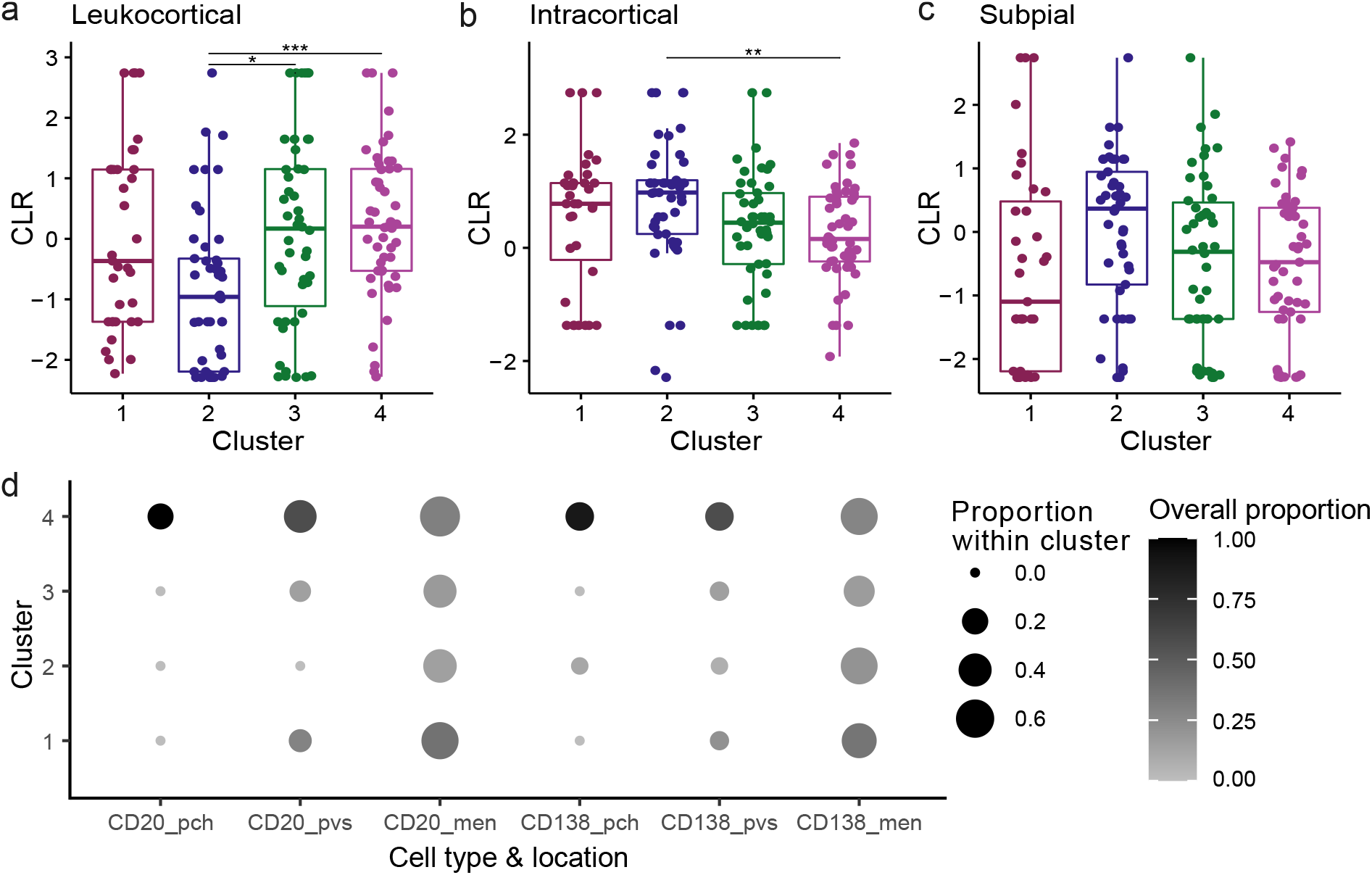
Cortical lesion proportions and B cell presence per MS subgroup. **a-c** Box plots showing the transformed proportions of leukocortical, intracortical and subpial lesions for 175 donors (cluster 1: n = 35; 2: n = 45; 3: n = 46; 4: n = 49). Significance was assessed by pairwise Mann-Whitney tests and adjusted for multiple testing (Bonferroni). *p ≤ 0.05; **p < 0.01; ***p < 0.001; ****p < 0.0001. **d** Dot plot displaying the proportion of B cell-positive donors per cluster (dot size) and the distribution of B cell-positive donors over the clusters (dot color). B cell presence/absence was investigated for 132 donors (1: n = 47; 2: n = 24; 3: n = 30; 4: n = 31). As an example, CD20^+^ B cells were found in the brain parenchyma of 6 donors. Of these 6 positive donors, all were part of cluster 4, resulting in an ‘overall’ proportion of 6/6 = 1, as indicated by the black dot in the first column. The within cluster proportion is 6/31 = 0.19, as indicated by dot size. Significance was assessed by Fisher’s exact test per combination of location and cell type, followed by pairwise Fisher’s exact tests adjusted for multiple testing to compare the clusters. P-values are reported in the accompanying text. CLR = centered log ratio; pch = parenchyma; pvs = perivascular space; men = meninges

The manifestation and frequency of clinical signs and symptoms is largely similar for the MS donors of the different subgroups (Fig. 3a). In line with the faster disease progression and earlier death of donors in cluster 4, temporal data shows that donors in cluster 4 generally experience motor, sensory and other symptoms earlier than those in the other subgroups (Fig. 3b), even though age at onset is comparable. Thus, it is the timing rather than the type of signs and symptoms that distinguishes the subgroups and identifies donors in cluster 4 as those most severely affected by MS.

### Cluster 2 has a different pattern of cortical lesion pathology

The MS subgroups are mainly based on white matter neuropathology; only one binary variable in the input dataset was related to the presence of cortical lesions. To further investigate cortical pathology, it was determined whether the subgroups differ with regards to the locations of cortical lesions. The proportion of leukocortical lesions is generally lower in donors of cluster 2 (2 vs 3: p = 0.015; 2 vs 4: p = 1.7 × 10^−4^; Fig. 4a), while the proportion of intracortical lesions is higher (2 vs 4: p = 0.007; Fig. 4b). Although cluster 2 also has the highest proportion of subpial lesions, the difference between the subgroups is not significant (p = 0.052; Fig. 4c). The number of cortical lesions identified is lowest for cluster 1 and highest for cluster 4, but this difference is not significant when correcting for the number of cortical tissue blocks investigated (suppl. Fig. 5a,b; Online Resource 3). Therefore, it seems that there is no marked difference in cortical lesion load when considering only those donors with one or more cortical lesions. The pattern of cortical lesions does differ for cluster 2, signifying a relation between white and gray matter pathology in MS.

**Fig. 5.**
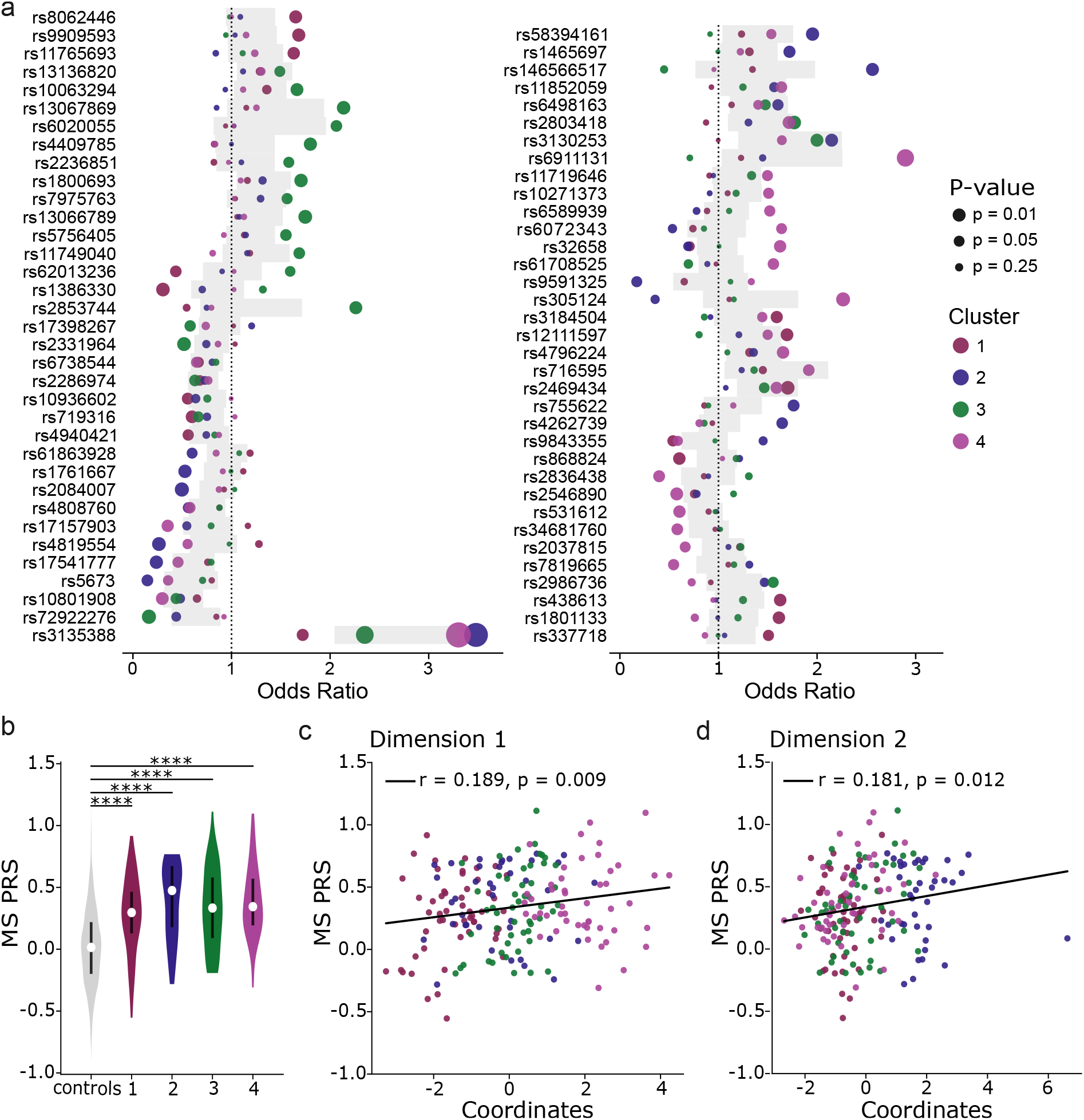
Genetic analysis of the MS subgroups and dimensions. Genetic data was available for 192 MS donors (cluster 1: n = 53; 2: n = 39; 3: n = 51; 4: n = 49) and 2049 controls. **a** Plot showing the odds ratios of the 70 MS-associated variants with an unadjusted p-value < 0.05 for at least one of the subgroups. The color of the dot indicates the cluster; dot size reflects the unadjusted p-value. The gray shaded area represents the 95% confidence interval of the odds ratio for the comparison between donors of all four MS subgroups as cases and non-MS non-NMO donors as controls. **b** Violin plot showing the MS PRS for the controls and MS subgroups. The white dot in the body of the violin indicates the median, the black line the interquartile range. Significance was assessed by pairwise Mann-Whitney tests and adjusted for multiple testing (Bonferroni). *p ≤ 0.05; **p < 0.01; ***p < 0.001; ****p < 0.0001. **c, d** Scatter plots of MS PRS against the coordinates of donors on dimension 1 (**c**) and 2 (**d**). The fitted regression line, Pearson correlation coefficient (r) and p-value (Wald Test with t-distribution of the test statistic) are shown. The color of the dots indicates the cluster, following the same scheme as in **a**. For plots of the PRS against the coordinates of donors on dimension 3-5, see suppl. Fig. 6. PRS = polygenic risk score

### The proportion of donors with B cells in the brainstem is higher in cluster 4

Studying the same autopsy cohort, Fransen et al. [17] found that an absence of B cells at specific anatomical locations was associated with clinically and pathologically less severe MS. This allowed us to determine the distribution of donors with and without B cells at these locations over the subgroups (Fig. 4d). Previously, it was observed that CD20^+^ B cells and CD138^+^ plasma cells were only infrequently present at various brainstem locations, except for the meninges [17]. Correspondingly, the proportion of donors with B cells in the meninges is relatively constant across all four subgroups (CD20^+^ cells: p = 0.12; CD138^+^ cells: p = 0.33). However, the subgroups differ with regards to the proportions of donors with B cells in the brain parenchyma (CD20^+^ cells: p = 2.2 × 10^−4^; CD138^+^ cells: p = 3.7 × 10^−5^) and perivascular space (CD20^+^ cells: p = 6.0 × 10^−4^; CD138^+^ cells: p = 0.037). Cluster 4 has relatively more B cell-positive donors compared to the other subgroups (for CD20^+^ cells in the parenchyma: 1 vs 4 (p = 0.017); CD138^+^ cells in the parenchyma: 1 vs 4 (p = 0.002) and 3 vs 4 (p = 0.028), and CD20^+^ cells in the perivascular space: 2 vs 4 (p = 0.003)).

Importantly, Fransen et al. [17] found that B cells were more often observed in brainstem tissue blocks with lesions as compared to those without lesions. The subgroups differ significantly in this aspect (p = 2.2 × 10^−5^; suppl. Fig. 5c, Online Resource 3). The presence of B cells was more often investigated in blocks with a lesion for both cluster 3 and 4 compared to cluster 1 and 2. Since cluster 4, but not cluster 3, has a higher proportion of donors with B cells at various locations, this suggests that cluster 4 could be characterized by an increased presence of B cells independent of lesion presence. Accordingly, this is also the subgroup showing the most profound presence of perivascular cuffing (suppl. Fig. 3d, Online Resource 3), supporting the association of immune cell infiltration with the most severe form of MS pathology.

### Associations with MS genetic risk variants and polygenic risk score corroborate the subgroups

Each subgroup was compared with non-MS non-NMO controls for MS-associated variants, to evaluate whether the clustering analysis was able to reduce heterogeneity [44]. Of the 288 variants that were investigated, 70 are putatively associated with one or more subgroups (unadjusted p-value < 0.05; Fig. 5a). Despite the small sample size, one association remains significant after correction for 4 × 288 tests: rs3135388A with cluster 2 and 4 (controls vs 2: p = 3.6 × 10^−8^; vs 4: p = 6.1 × 10^−9^ (unadjusted p-values)). This SNP is highly correlated with the HLA-DRB1*1501 allele [10, 21], the strongest genetic risk factor for MS. Furthermore, Fig. 5a shows that the odds ratios are frequently extreme for one of the subgroups, falling outside the 95% confidence interval of the odds ratio for the comparison between all MS donors combined and the controls (i.e. the gray shaded area). This could indicate a potential genetic basis for the neuropathological subgroups [44].

To further investigate the genetics of the subgroups, MS PRSs were calculated. PRSs are higher for the MS subgroups compared to controls (controls vs 1: p = 1.1 × 10^−7^; vs 2: p = 2.4 × 10^−9^; vs 3: p = × 10^−9^; vs 4: p = 5.1 × 10^−13^; Fig. 5b), but do not differ significantly between subgroups. Importantly, the MS PRS does correlate significantly with the coordinates of the MS donors on the first and second dimension (Fig. 5c,d), but not with dimension 3-5 (suppl. Fig. 6). Dimension 1 and 2 each explain around 3.5% of the variance in the MS PRS (R^2^ of 0.036 and 0.033, respectively), lending further genetic support for the current subdivision.

## Discussion

To improve the prediction of disease course and choice of disease-modifying therapies, a subdivision of MS based on pathobiological mechanisms is of key importance. Using a data-driven clustering approach, we identified four distinct subgroups of MS brain donors based on post-mortem white matter neuropathologic characteristics. Our subgroups correspond to differences on the clinical and genetic level and are associated with distinct cortical and B cell pathology. To what extent these subgroups are qualitatively different and/or represent multiple axes along which patients vary remains to be determined.

Cluster 1 is characterized by a low lesion load and a high proportion of inactive and remyelinated lesions. Combined with a median time from disease onset to EDSS-6 of 17 years, this subgroup reflects a comparatively mild form of MS. Previous research found that patients with benign MS (commonly defined as EDSS ≤ 3.0 after ≥ 15 years from disease onset [8]) have a significantly lower degree of cortical pathology on MRI [7], which appears to be in line with the lower prevalence of cortical lesions in cluster 1. Benign MS has also been associated with a younger age at onset and female sex [8]. However, no significant differences between the subgroups in age at onset or the proportion of females were observed. Especially the latter was contrary to expectations, since sex-specific differences in MS have been described. Clinically, females are characterized by a higher relapse rate (indicating more inflammatory activity), while males accumulate more disability (which has been related to a more predominant neurodegenerative component of the disease) [37]. Neuropathologically, males have a higher proportion of mixed lesions and more often cortical pathology than females [18, 35]; this difference was observed in the same NBB cohort studied here [35]. The current analysis seems to suggest that differences between the sexes become less apparent when considering a combination of different neuropathological variables combined (i.e. the subgroups), compared to individual lesion types.

MS donors in cluster 2 score high on the second dimension, which is related to increased numbers of reactive sites, the presence of nodules and a high proportion of active lesions with ramified microglia. In general, the proportion of active lesions is low in patients with a long disease duration [18, 35]; patients with progressive MS with attacks were found to have a higher proportion of active lesions than those without clinical attacks [18]. This raises the possibility that donors in cluster 2 had a different disease experience than those in the other subgroups. Although the current temporal clinical analysis showed that donors in cluster 2 generally experienced signs and symptoms at a later age, the resolution is too low to distinguish between MS attacks and disease-free periods. Furthermore, the co-occurrence of nodules, accumulations of HLA^+^ cells in normal-appearing white matter, and active lesions has been described previously [54]. These nodules have been considered ‘pre-active’ lesions, although it was recognized that most would probably not progress into demyelinated lesions [11, 54]. Alternatively, a study on biopsy tissue from early MS patients found that nodules form in response to axonal transection in lesions and subsequent Wallerian degeneration [51]. Although the meaning of these sites is unclear, it is interesting that both studies found signs of pro- and anti-inflammatory microglia phenotypes, with inter-individual differences [51, 54]. This allows for the possibility that the subgroups differ with regards to the role of microglia accumulations in the disease process, despite the lack of significant quantitative differences between clusters 2, 3 and 4. In this regard, it is interesting that the HLA-DRB1*1501 risk allele, most strongly associated with cluster 2 and 4, has been shown to affect microglial inflammation [57].

In addition, cluster 2 seems to have a different pattern of cortical lesions, with relatively less leukocortical and more intracortical lesions. This could be related to the comparatively lower proportion of mixed lesions in cluster 2, since mixed lesions have previously been associated with leukocortical and intracortical lesions (but not subpial lesions) in this autopsy cohort [35]. A recent MRI study also notes this association between leukocortical and mixed (paramagnetic rim) lesions [3]. In contrast, subpial lesion burden was less strongly related to white matter lesion volumes, possibly indicating (partially) different mechanisms [3]. Correspondingly, our MS subgroups based on white matter pathology do not differ significantly with regards to subpial lesions.

Cluster 1 and 2 both have a high proportion of remyelinated lesions relative to cluster 3 and 4. Previously, extensive remyelination has been observed in around one fifth of MS patients, which was associated with older age at death and longer disease duration (but not with sex or age at onset) [46]. This seems to fit with the donors in cluster 1 and 2, who indeed died at an older age, after a longer period of disease. In contrast, cluster 3 and 4 both have relatively few remyelinated lesions. This remyelination failure can be attributed to several factors, such as a hostile tissue environment with pro-inflammatory myeloid cells, reduced clearance of myelin debris, and/or a problem with myelin production or oligodendrocyte maturation [4, 14, 22, 30]. The difference in the proportion of inactive lesions between cluster 3 and cluster 4, combined with the distinct microglia morphology characterizing these subgroups, might hint at different mechanisms of remyelination failure.

Clinically and pathologically, cluster 4 has the most severe form of MS. The proportion of donors with B cells in the brainstem was highest for this subgroup, in accordance with prior post-mortem research relating B cells to worse clinical outcomes [43]. Moreover, perivascular B cells were associated with higher levels of T cells and macrophages/microglia [43], which seems to fit with the relatively high proportion of donors with leukocyte cuffs and the predominance of lesions populated with foamy microglia in cluster 4.

Some caution is necessary when interpreting the subgroups, because of limitations inherent to the data-driven approach and the dataset. For instance, while the selection of the first five dimensions removed redundancy and noise, relevant data may have been discarded. Conversely, it is possible that some dimensions - especially dimension 4 and 5, which explain less variation and are more difficult to interpret - have contributed noise to the clusters. In this regard, it is important to note that the first two dimensions correlate with the MS PRS, indicating that these might give insight into MS neuropathogenicity. This also gives credibility to the current subdivision, because cluster 1, 2 and 4 score distinctly on these dimensions.

However, a lack of correlation with the PRS does not necessarily mean that the dimension is irrelevant for clustering. Whereas the PRS reflects genetic susceptibility to MS onset, it may capture disease progression less well, since this has been associated with distinct genetic factors and pathological processes [15, 24]. For instance, the MHC region is clearly associated with disease onset [21, 47], but not related to clinical disability as measured by the MS severity score [24]. In this regard, it is notable that the HLA-DRB1*1501 risk allele is strongly associated with two out of four subgroups, in line with previous reports associating this allele with post-mortem neuropathology [56, 57]. This argues for a correlation between the onset of MS and the pathology observed at autopsy and is indicative of the complex relation between MS susceptibility, clinical course and neuropathological processes.

Furthermore, the subgroups may partially reflect differences in MS treatment and autopsy procedure. Donors in cluster 2 died more recently and would have more often been dissected under guidance of MRI, which has been associated with increased detection of reactive sites and active lesions [11]. This might have affected the distinction between cluster 1 and 2; the difference between cluster 2, 3 and 4 is more robust, since mixed lesions are detected relatively well even without the use of MRI [11]. In addition, donors in cluster 2 may have had more disease-modifying treatment options available during their lifetime. Since this subgroup is characterized by relatively mild MS, it is tempting to speculate that more recent therapeutic options have had an effect on neuropathology and that their use is reflected in the composition of the subgroups. Importantly, a historical trend towards less severe MS has amongst others been related to the emergence of effective disease-modifying therapies [27].

It remains unclear to what extent the subgroups are explained by different disease durations and age-related changes, gradations in disease severity and/or biologically coherent subtypes, in part because of the cross-sectional nature of the data. It is possible that the different lesion and immune cell patterns are mainly a reflection of differences in disease duration and severity between the subgroups. Alternatively, this study offers the intriguing possibility of at least two neuropathogenetic mechanisms in MS (reflected in the dimensions) that are more or less ‘active’ in a certain individual. This is similar to the view of Kuhlmann et al. [29], who suggest that instead of one disease mechanism underlying MS, there is a combination of mechanisms of injury and repair, with varying importance between patients and over time. For instance, dimension 1 seems to represent a combination of non-resolving inflammation and failure of remyelination, which appears to drive disease progression in cluster 4 but is less active in cluster 1 and 2; dimension 2 could be related to a specific form of microglial (re)activity, mostly relevant for donors in cluster 2. Importantly, our exploratory genetic analysis could support the existence of distinct biological subtypes, although it was underpowered to identify genetic associations and further investigation is necessary. The HLA-DRB1*1501 tagging SNP is an exception, possibly related to its large effect size and the role of HLA-staining in the characterization of lesions.

Our findings exemplify the importance of data-driven analysis on large and well-characterized autopsy cohorts. This and other data-driven studies give insight into MS neuropathology unbiased by *a priori* hypotheses and can help with in-depth characterization of disease mechanisms and their activity in individual patients. Future studies should aim to incorporate an even larger number of samples, and more extensive information regarding neuropathology and clinical background (including drug usage). Importantly, the neuropathology-based subgroups were not distinguishable based on clinical signs and symptoms alone, and only partially based on disease trajectory. Thus, achieving stratification of MS patients based on pathobiology in clinical and research settings will also require a firm link between these neuropathological mechanisms and biomarkers - including but not limited to genetic and imaging factors.

## Data Availability

All data produced in the present study are available upon reasonable request to the authors.

## Acknowledgements

We are grateful to those who donated their brain for research to the Netherlands Brain Bank. We would like to acknowledge the ‘Vrienden van het Herseninstituut’ for their contributions to this project. Figure 1a was created with BioRender.com.

