## Supplementary material for "Identification of neuropathology-based subgroups in multiple sclerosis using a data-driven approach": suppl. Table 1 and 2

**Online Resource 1: suppl. Table 1 and 2, related to the genetic analysis**

**Suppl. Table 1** Description of the 2049 non-MS non-NMO NBB donors that served as controls for the genetic analyses. MS = multiple sclerosis, NMO = neuromyelitis optica, NBB = Netherlands Brain Bank, SD = standard deviation; p = percentile

| **Age (years)** | | | **Year of death** | | | **Gender** | |
|---|---|---|---|---|---|---|---|
| **Mean (SD)** | **Median (p25-p75)** | **Range (min-max)** | **Mean (SD)** | **Median (p25-p75)** | **Range (min-max)** | **Males (%)** | **Females (%)** |
| 76.4 (12.14) | 78 (69 – 85) | 20 – 103 | 2006.88 (8.03) | 2008 (2000 – 2014) | 1988 – 2020 | 908 (44.31%) | 1141 (55.69%) |

**Suppl. Table 2** Description of the 288 MS-associated variants included. Chromosome number and genomic position are given in the column 'Chrom:Pos' (Genome Reference Consortium Human Build 37/hg19). The A1 and A2 alleles are presented as reported by PLINK [5], with A1 being the allele for which the odds ratio was calculated. N = 180 risk variants were identified in the genome-wide association study (GWAS) by Patsopoulos et al [4]; n = 9 single nucleotide polymorphisms (SNPs) were in strong linkage disequilibrium (LD) with an MS GWAS risk variant. N = 99 variants were related to MS pathology or severity, as identified by Fransen et al [2]; n = 1 SNP was in strong LD with a Fransen indel variant (rs333). Two variants were identified by both studies. Finally, the tagging SNP of the HLA-DRB1*15:01 allele was included [1, 3]. Note that the rs number provided here may not always correspond to the one reported earlier

| **Rs number** | **Chrom:Pos** | **A1** | **A2** | **Reason for inclusion** |
|---|---|---|---|---|
| rs6670198 | 1:2520527 | C | T | GWAS |
| rs2986736 | 1:6512547 | C | T | GWAS |
| rs1801133 | 1:11856378 | A | G | GWAS |
| rs2236851 | 1:25243554 | T | C | Fransen et al, pathology |
| rs6672420 | 1:25291010 | T | A | GWAS |
| rs79979643 | 1:32738415 | A | G | GWAS |
| rs12127450 | 1:49067014 | C | T | Fransen et al, severity |
| rs72922276 | 1:65429319 | A | G | GWAS |
| rs17541777 | 1:71375500 | C | T | Fransen et al, severity |
| rs5673 | 1:71477841 | T | A | Fransen et al, severity |
| rs11161550 | 1:85682020 | A | G | GWAS |
| rs35486093 | 1:85729820 | G | A | GWAS |
| rs12133753 | 1:92222089 | T | C | GWAS |
| rs58394161 | 1:92939959 | C | T | GWAS |
| rs11809700 | 1:93152635 | T | C | GWAS |
| rs6681429 | 1:93427406 | C | A | In strong LD with rs1415069 (GWAS) |
| rs74449127 | 1:101290432 | G | A | GWAS |
| rs11578655 | 1:101412902 | G | T | GWAS |
| rs17505688 | 1:107699041 | C | T | Fransen et al, severity |
| rs10801908 | 1:117090493 | T | C | GWAS |
| rs483180 | 1:120267505 | G | C | GWAS |
| rs3014866 | 1:153329071 | T | C | Fransen et al, pathology |
| rs112344141 | 1:154983036 | G | T | GWAS |

| | | | | |
|---|---|---|---|---|
| rs2317231 | 1:157686337 | T | G | GWAS |
| rs3737798 | 1:160389984 | G | A | GWAS |
| rs6427540 | 1:160634588 | T | C | GWAS |
| rs983494 | 1:160703965 | A | G | GWAS |
| rs1323292 | 1:192541021 | G | A | GWAS |
| rs1061170 | 1:196659237 | C | T | Fransen et al, pathology |
| rs59655222 | 1:200875897 | C | T | GWAS |
| rs2796267 | 1:207924906 | G | A | Fransen et al, pathology |
| rs9308424 | 1:212877776 | A | G | GWAS |
| rs13387792 | 2:1912219 | A | G | Fransen et al, severity |
| rs1869410 | 2:5257356 | C | T | Fransen et al, severity |
| rs11899404 | 2:12607893 | T | C | GWAS |
| rs11125803 | 2:25052177 | C | T | GWAS |
| rs10178552 | 2:25973309 | T | C | Fransen et al, severity |
| rs12478539 | 2:43355324 | C | G | GWAS |
| rs17398267 | 2:48937823 | G | T | Fransen et al, severity |
| rs13019537 | 2:48976577 | G | C | Fransen et al, severity |
| rs1177228 | 2:61242410 | A | G | GWAS |
| rs13385171 | 2:65661843 | T | C | GWAS |
| rs12622670 | 2:68646536 | C | T | GWAS |
| rs72823440 | 2:112490418 | A | G | In strong LD with rs71252597 (GWAS) |
| rs57116599 | 2:112770799 | A | G | GWAS |
| rs423904 | 2:113887262 | T | C | Fransen et al, severity |
| rs3814022 | 2:135047919 | G | C | Fransen et al, severity |
| rs10191360 | 2:136884679 | T | C | GWAS |
| rs962052 | 2:151644203 | C | T | GWAS |
| rs6738544 | 2:191989356 | A | C | GWAS |
| rs2037815 | 2:202101715 | G | A | Fransen et al, pathology |
| rs3116496 | 2:204594512 | C | T | Fransen et al, pathology |
| rs5742909 | 2:204732347 | T | C | Fransen et al, pathology |
| rs231775 | 2:204732714 | G | A | Fransen et al, pathology |
| rs35540610 | 2:231121829 | C | T | GWAS |
| rs1597944 | 2:234504098 | C | T | Fransen et al, severity |
| rs9863496 | 3:18798848 | C | T | GWAS |
| rs13327021 | 3:27783015 | T | C | GWAS |
| rs438613 | 3:28072086 | C | T | GWAS |
| rs11919880 | 3:32962051 | G | A | GWAS |
| rs113341849 | 3:46384204 | A | G | In strong LD with rs333 (Fransen et al, pathology) |
| rs1799987 | 3:46411935 | G | A | Fransen et al, pathology |
| rs11719646 | 3:56467691 | G | A | Fransen et al, severity |
| rs9878602 | 3:71535338 | G | T | GWAS |
| rs111430408 | 3:100848597 | T | C | GWAS |
| rs4325907 | 3:101749022 | C | T | GWAS |
| rs2289746 | 3:105455955 | T | C | GWAS |
| rs138433213 | 3:112693983 | G | T | GWAS |

| | | | | |
|---|---|---|---|---|
| rs9843355 | 3:119228508 | A | G | GWAS |
| rs2331964 | 3:121542898 | T | C | GWAS |
| rs75937181 | 3:121783015 | G | T | GWAS |
| rs1014486 | 3:159691112 | C | T | GWAS |
| rs10936602 | 3:169536637 | C | T | GWAS |
| rs13067869 | 3:173665677 | G | T | Fransen et al, severity |
| rs2590438 | 3:187565968 | G | T | GWAS |
| rs13066789 | 3:187987624 | C | T | GWAS |
| rs8192678 | 4:23815662 | T | C | Fransen et al, severity |
| rs305124 | 4:39769586 | G | A | Fransen et al, severity |
| rs13136820 | 4:40307564 | C | T | GWAS |
| rs6837324 | 4:48127262 | G | A | GWAS |
| rs2705616 | 4:87862396 | C | G | GWAS |
| rs2853744 | 4:88896248 | T | G | Fransen et al, severity |
| rs6533052 | 4:103911781 | A | G | GWAS |
| rs2726479 | 4:106255589 | T | C | GWAS |
| rs10516537 | 4:107628071 | A | C | Fransen et al, severity |
| rs9992763 | 4:109058718 | G | T | GWAS |
| rs17051321 | 4:122119449 | T | C | GWAS |
| rs2069762 | 4:123377980 | C | A | Fransen et al, severity |
| rs12644284 | 4:154154000 | G | A | Fransen et al, severity |
| rs72989863 | 4:164493807 | A | G | GWAS |
| rs34681760 | 5:6712834 | T | C | GWAS |
| rs11750073 | 5:11548050 | T | C | Fransen et al, severity |
| rs10078091 | 5:25495005 | A | G | Fransen et al, severity |
| rs10063294 | 5:35877505 | G | A | GWAS |
| rs11749040 | 5:40396425 | A | G | GWAS |
| rs6880809 | 5:40429250 | T | A | GWAS |
| rs7731626 | 5:55444683 | A | G | GWAS |
| rs32658 | 5:118703662 | T | G | GWAS |
| rs244656 | 5:133449827 | T | A | GWAS |
| rs2084007 | 5:133891282 | T | C | GWAS |
| rs2569190 | 5:140012916 | A | G | Fransen et al, pathology |
| rs249677 | 5:141539339 | C | A | GWAS |
| rs6198 | 5:142657621 | C | T | Fransen et al, pathology |
| rs41423247 | 5:142778575 | C | G | Fransen et al, pathology |
| rs6190 | 5:142780337 | T | C | Fransen et al, pathology |
| rs10052957 | 5:142786701 | A | G | Fransen et al, pathology |
| rs3212227 | 5:158742950 | G | T | Fransen et al, pathology |
| rs2546890 | 5:158759900 | G | A | GWAS |
| rs11957313 | 5:169950394 | A | G | Fransen et al, severity |
| rs67111717 | 5:176790162 | G | A | GWAS |
| rs13208234 | 6:7119134 | G | A | In strong LD with rs12211604 (GWAS) |
| rs111635774 | 6:14691215 | T | C | GWAS |
| rs6941421 | 6:15089151 | C | T | Fransen et al, severity |

| | | | | |
|---|---|---|---|---|
| rs719316 | 6:16672760 | C | T | GWAS |
| rs3130253 | 6:29634012 | A | G | Fransen et al, pathology |
| rs3135388 | 6:32413051 | A | G | tag for the HLA-DRB1*15:01 allele |
| rs1076928 | 6:36348689 | T | C | GWAS |
| rs72928038 | 6:90976768 | A | G | Fransen et al, pathology & GWAS |
| rs6899560 | 6:96275538 | G | A | Fransen et al, severity |
| rs9480865 | 6:108916573 | C | T | Fransen et al, severity |
| rs11542663 | 6:119215402 | C | A | GWAS |
| rs802730 | 6:128280104 | C | T | GWAS |
| rs75191738 | 6:130348257 | T | C | GWAS |
| rs2327586 | 6:135495226 | T | C | GWAS |
| rs58761508 | 6:135835901 | A | G | In strong LD with rs4896153 (GWAS) |
| rs62420820 | 6:137438057 | A | G | GWAS |
| rs631204 | 6:137959455 | A | C | GWAS |
| rs17780048 | 6:138179146 | T | C | GWAS |
| rs263153 | 6:142949309 | T | G | Fransen et al, severity |
| rs6911131 | 6:143865221 | G | A | GWAS |
| rs7744583 | 6:157197515 | A | G | Fransen et al, severity |
| rs1738074 | 6:159465977 | T | C | GWAS |
| rs12202350 | 6:160379096 | C | T | Fransen et al, severity |
| rs6917747 | 6:160402705 | A | G | Fransen et al, severity |
| rs6944568 | 7:2448317 | A | C | In strong LD with rs55858457 (GWAS) |
| rs10951042 | 7:3139417 | C | T | GWAS |
| rs156429 | 7:23306020 | C | T | Fransen et al, pathology |
| rs10951154 | 7:27135314 | C | T | GWAS |
| rs10245867 | 7:28142186 | T | G | GWAS |
| rs12111597 | 7:34870001 | A | G | Fransen et al, severity |
| rs60600003 | 7:37382465 | G | T | GWAS |
| rs10230723 | 7:50239880 | T | A | GWAS |
| rs116877451 | 7:50328339 | G | A | GWAS |
| rs11765693 | 7:75985373 | G | A | Fransen et al, severity |
| rs1761667 | 7:80244939 | G | A | Fransen et al, pathology |
| rs17157903 | 7:103628036 | T | C | Fransen et al, severity |
| rs73414214 | 7:105706462 | A | C | GWAS |
| rs868824 | 7:110391921 | C | T | Fransen et al, severity |
| rs10243024 | 7:116346603 | A | G | Fransen et al, severity |
| rs4728142 | 7:128573967 | A | G | GWAS |
| rs10271373 | 7:138729795 | A | C | GWAS |
| rs354033 | 7:149289464 | A | G | GWAS |
| rs6994992 | 8:31495581 | T | C | Fransen et al, pathology |
| rs2116078 | 8:73363989 | T | G | Fransen et al, severity |
| rs28703878 | 8:79417222 | G | A | GWAS |
| rs78727559 | 8:95851818 | G | T | GWAS |
| rs10505082 | 8:106780628 | A | G | Fransen et al, severity |
| rs735542 | 8:128175696 | G | A | GWAS |

| | | | | |
|---|---|---|---|---|
| rs6990534 | 8:128814091 | A | G | GWAS |
| rs7819665 | 8:129177769 | C | T | GWAS |
| rs3923387 | 8:144986793 | T | C | GWAS |
| rs16925027 | 9:6978321 | G | A | Fransen et al, severity |
| rs10977017 | 9:8380546 | A | G | Fransen et al, severity |
| rs2803418 | 9:78909274 | T | G | Fransen et al, severity |
| rs7855251 | 9:100868189 | C | T | GWAS |
| rs4880213 | 9:140031001 | T | C | Fransen et al, pathology |
| rs12722559 | 10:6070273 | A | C | GWAS |
| rs11256593 | 10:6117322 | C | T | GWAS |
| rs1399180 | 10:8098719 | T | C | GWAS |
| rs2399849 | 10:12499860 | A | G | Fransen et al, severity |
| rs1927457 | 10:30008663 | C | T | Fransen et al, severity |
| rs793102 | 10:31391564 | T | C | In strong LD with rs1087056 (GWAS) |
| rs61863928 | 10:64449549 | T | G | GWAS |
| rs4747075 | 10:72449419 | A | G | Fransen et al, severity |
| rs17741873 | 10:75653800 | T | G | GWAS |
| rs1250551 | 10:81059335 | T | G | GWAS |
| rs1800682 | 10:90749963 | G | A | Fransen et al, pathology |
| rs2234978 | 10:90771829 | T | C | Fransen et al, severity |
| rs1112718 | 10:94479107 | G | A | GWAS |
| rs716595 | 10:112006486 | A | G | Fransen et al, severity |
| rs2766051 | 10:129121892 | A | G | Fransen et al, severity |
| rs7914524 | 10:130104867 | T | C | Fransen et al, severity |
| rs61884005 | 11:14402930 | G | C | GWAS |
| rs117361591 | 11:14861957 | T | G | In strong LD with rs570429157 (GWAS) |
| rs1365120 | 11:36438075 | C | T | GWAS |
| rs2269434 | 11:47360412 | C | T | GWAS |
| rs4939490 | 11:60793651 | G | C | GWAS |
| rs11231749 | 11:64095178 | C | T | GWAS |
| rs531612 | 11:65705432 | C | T | GWAS |
| rs1386330 | 11:87819427 | C | T | Fransen et al, severity |
| rs4409785 | 11:95311422 | C | T | GWAS |
| rs56095240 | 11:95421830 | A | T | GWAS |
| rs34026809 | 11:118480695 | C | G | GWAS |
| rs12365699 | 11:118743286 | A | G | GWAS |
| rs6589706 | 11:118747813 | A | G | GWAS |
| rs149114341 | 11:118783424 | A | G | GWAS |
| rs6589939 | 11:122518525 | G | A | GWAS |
| rs4262739 | 11:128421175 | A | G | GWAS |
| rs1800693 | 12:6440009 | C | T | GWAS |
| rs2364485 | 12:6514963 | A | C | GWAS |
| rs7977720 | 12:9866349 | T | C | GWAS |
| rs261902 | 12:32476727 | A | G | Fransen et al, severity |
| rs701006 | 12:58106836 | A | G | GWAS |

| | | | | |
|---|---|---|---|---|
| rs2069727 | 12:68548223 | C | T | Fransen et al, pathology |
| rs61708525 | 12:94661453 | G | A | GWAS |
| rs3184504 | 12:111884608 | T | C | GWAS |
| rs7134248 | 12:121897052 | T | C | Fransen et al, severity |
| rs7975763 | 12:123604053 | T | C | GWAS |
| rs9591325 | 13:50811220 | C | T | GWAS |
| rs9568402 | 13:50961957 | T | A | GWAS |
| rs9319189 | 13:86618098 | A | G | Fransen et al, severity |
| rs77654077 | 13:100026952 | C | A | GWAS |
| rs2039485 | 14:32353250 | C | T | Fransen et al, severity |
| rs11852059 | 14:52306091 | C | A | GWAS |
| rs12434551 | 14:69253364 | T | A | GWAS |
| rs34695601 | 14:76014298 | C | T | GWAS |
| rs116899835 | 14:88523488 | T | C | GWAS |
| rs12588969 | 14:103230758 | G | C | GWAS |
| rs12147246 | 14:103265844 | A | G | GWAS |
| rs62013236 | 15:79247482 | T | C | GWAS |
| rs6496663 | 15:90887584 | C | A | GWAS |
| rs752092 | 15:101781934 | G | A | Fransen et al, severity |
| rs405343 | 16:1067832 | T | G | GWAS |
| rs1448239 | 16:10187435 | C | G | Fransen et al, severity |
| rs2286974 | 16:11114512 | G | A | GWAS |
| rs6498163 | 16:11213951 | C | T | GWAS |
| rs146566517 | 16:11353879 | T | C | GWAS |
| rs34947566 | 16:11412926 | A | C | GWAS |
| rs3809627 | 16:30103160 | A | C | GWAS |
| rs8062446 | 16:57077094 | T | C | GWAS |
| rs12925972 | 16:79111297 | T | C | GWAS |
| rs17724508 | 16:79350204 | C | T | GWAS |
| rs404694 | 16:79582798 | C | A | Fransen et al, severity |
| rs6564681 | 16:79652720 | C | T | GWAS |
| rs35703946 | 16:86021505 | A | G | GWAS |
| rs11652878 | 17:3654980 | G | A | Fransen et al, pathology |
| rs7211577 | 17:14114280 | G | A | Fransen et al, severity |
| rs1137933 | 17:26105932 | A | G | Fransen et al, severity |
| rs9892479 | 17:31421901 | T | G | Fransen et al, severity |
| rs1133763 | 17:32647831 | C | A | Fransen et al, pathology |
| rs2107538 | 17:34207780 | T | C | Fransen et al, pathology |
| rs4796224 | 17:34842521 | G | A | GWAS |
| rs876493 | 17:37824545 | G | A | Fransen et al, severity |
| rs9909593 | 17:37970149 | G | A | GWAS |
| rs883871 | 17:38252660 | A | G | GWAS |
| rs744166 | 17:40514201 | G | A | Fransen et al, pathology |
| rs11079784 | 17:45702280 | T | C | GWAS |
| rs2150879 | 17:57859210 | G | A | GWAS |

| | | | | |
|---|---|---|---|---|
| rs1318 | 17:65691382 | G | A | Fransen et al, severity |
| rs9900529 | 17:73335776 | C | G | GWAS |
| rs2028455 | 18:47966005 | T | C | Fransen et al, severity |
| rs1557351 | 18:54752314 | C | T | Fransen et al, severity |
| rs4940730 | 18:56269737 | G | A | GWAS |
| rs4940421 | 18:56359234 | G | C | In strong LD with rs9955954 (GWAS) |
| rs2469434 | 18:67544046 | C | T | GWAS |
| rs337718 | 18:69774278 | T | C | Fransen et al, severity |
| rs2074897 | 19:1391235 | A | G | Fransen et al, severity |
| rs12981030 | 19:4466486 | G | T | In strong LD with rs12971909 (GWAS) |
| rs1077667 | 19:6668972 | T | C | GWAS |
| rs34536443 | 19:10463118 | C | G | GWAS |
| rs28834106 | 19:10592144 | C | T | GWAS |
| rs12609500 | 19:11173928 | T | C | GWAS |
| rs7253363 | 19:11682495 | T | G | Fransen et al, severity |
| rs58166386 | 19:16559421 | G | A | GWAS |
| rs11666377 | 19:17118433 | T | C | Fransen et al, severity |
| rs4808760 | 19:18301979 | G | C | GWAS |
| rs1064395 | 19:19361735 | A | G | Fransen et al, pathology |
| rs7260482 | 19:45143942 | C | A | GWAS |
| rs11083862 | 19:47638539 | A | T | GWAS |
| rs1465697 | 19:49837246 | T | C | GWAS |
| rs3865444 | 19:51727962 | A | C | Fransen et al, pathology |
| rs299175 | 19:56313528 | A | G | Fransen et al, severity |
| rs6072343 | 20:39968188 | A | G | GWAS |
| rs4812772 | 20:42579051 | T | C | GWAS |
| rs6032662 | 20:44734310 | C | T | GWAS |
| rs6020055 | 20:48422095 | G | A | GWAS |
| rs2585447 | 20:52744437 | C | T | GWAS |
| rs2248137 | 20:52789743 | G | C | GWAS |
| rs6742 | 20:62374441 | T | C | GWAS |
| rs9808753 | 21:34787312 | G | A | Fransen et al, pathology & GWAS |
| rs2836438 | 21:39864727 | A | G | GWAS |
| rs4819554 | 22:17565035 | G | A | Fransen et al, pathology |
| rs9610458 | 22:22205353 | C | T | GWAS |
| rs755622 | 22:24236392 | C | G | Fransen et al, pathology |
| rs760517 | 22:37258986 | T | C | GWAS |
| rs5756405 | 22:37310954 | A | G | GWAS |
| rs137955 | 22:40291807 | T | C | GWAS |
| rs140522 | 22:50971266 | T | C | GWAS |
