## Supplementary material for "Identification of neuropathology-based subgroups in multiple sclerosis using a data-driven approach": suppl. Table 3 and 4

**Online Resource 2: suppl. Table 3 and 4**

**Suppl. Table 3** Description of the FAMD dimensions with eigenvalue > 1. FAMD = Factor Analysis of Mixed Data

| Dimension | Eigenvalue | Variance (%) | Cumulative variance (%) |
|---|---|---|---|
| **1** | 2.38 | 18.46 | 18.46 |
| **2** | 1.71 | 13.24 | 31.69 |
| **3** | 1.55 | 12.01 | 43.70 |
| **4** | 1.26 | 9.76 | 53.46 |
| **5** | 1.12 | 8.70 | 62.17 |

**Suppl. Table 4** Autopsy characteristics of the MS subgroups. The column 'n' indicates the number of donors for whom the variable is known. SD = standard deviation; p = percentile

| | **pH** | | | | **Post-mortem delay (hours)** | | | |
|---|---|---|---|---|---|---|---|---|
| **Cluster** | **n** | **Mean (SD)** | **Median (p25 – p75)** | **Range (min – max)** | **n** | **Mean (SD)** | **Median (p25 – p75)** | **Range (min – max)** |
| 1 (n = 63) | 59 | 6.48 (0.29) | 6.46 (6.32 – 6.64) | 5.77 – 7.18 | 63 | 8.60 (4.97) | 7.92 (6.83 – 9.08) | 4.13 – 41.0 |
| 2 (n = 52) | 41 | 6.45 (0.22) | 6.44 (6.32 – 6.60) | 5.92 – 6.90 | 44 | 9.13 (5.42) | 8.75 (7.08 – 10.02) | 4.58 – 42.0 |
| 3 (n = 56) | 44 | 6.56 (0.45) | 6.44 (6.29 – 6.75) | 5.80 – 8.20 | 52 | 9.42 94.67) | 9.08 (7.06 – 9.85) | 3.00 – 35.0 |
| 4 (n = 57) | 53 | 6.42 (0.30) | 6.47 (6.25 – 6.62) | 5.68 – 7.22 | 56 | 9.30 | 8.50 (7.19 – 10.27) | 3.67 – 52.5 |

| | **Brain weight (grams)** | | | |
|---|---|---|---|---|
| **Cluster** | **n** | **Mean (SD)** | **Median (p25 – p75)** | **Range (min – max)** |
| 1 (n = 63) | 61 | 1188 (117) | 1200 (1101 – 1278) | 904 – 1413 |
| 2 (n = 52) | 44 | 1188 (147) | 1169 (1084 – 1289) | 940 – 1583 |
| 3 (n = 56) | 51 | 1195 (145) | 1190 (1114 – 1290) | 862 – 1475 |
| 4 (n = 57) | 55 | 1199 (128) | 1182 (1103 – 1293) | 969 – 1462 |
