## Supplementary material for "Identification of neuropathology-based subgroups in multiple sclerosis using a data-driven approach": suppl. Fig. 1 - 6

**Online Resource 3: suppl. Figure 1 – 6**

**Suppl. Figure 1** Factor maps with the MS donors (n = 228) plotted on dimension 2 and 3 (**a**), 3 and 4 (**b**), and 4 and 5 (**c**). Color and point shape indicate the cluster a donor belongs to. The explained variance of the dimension is given between brackets in the axis label

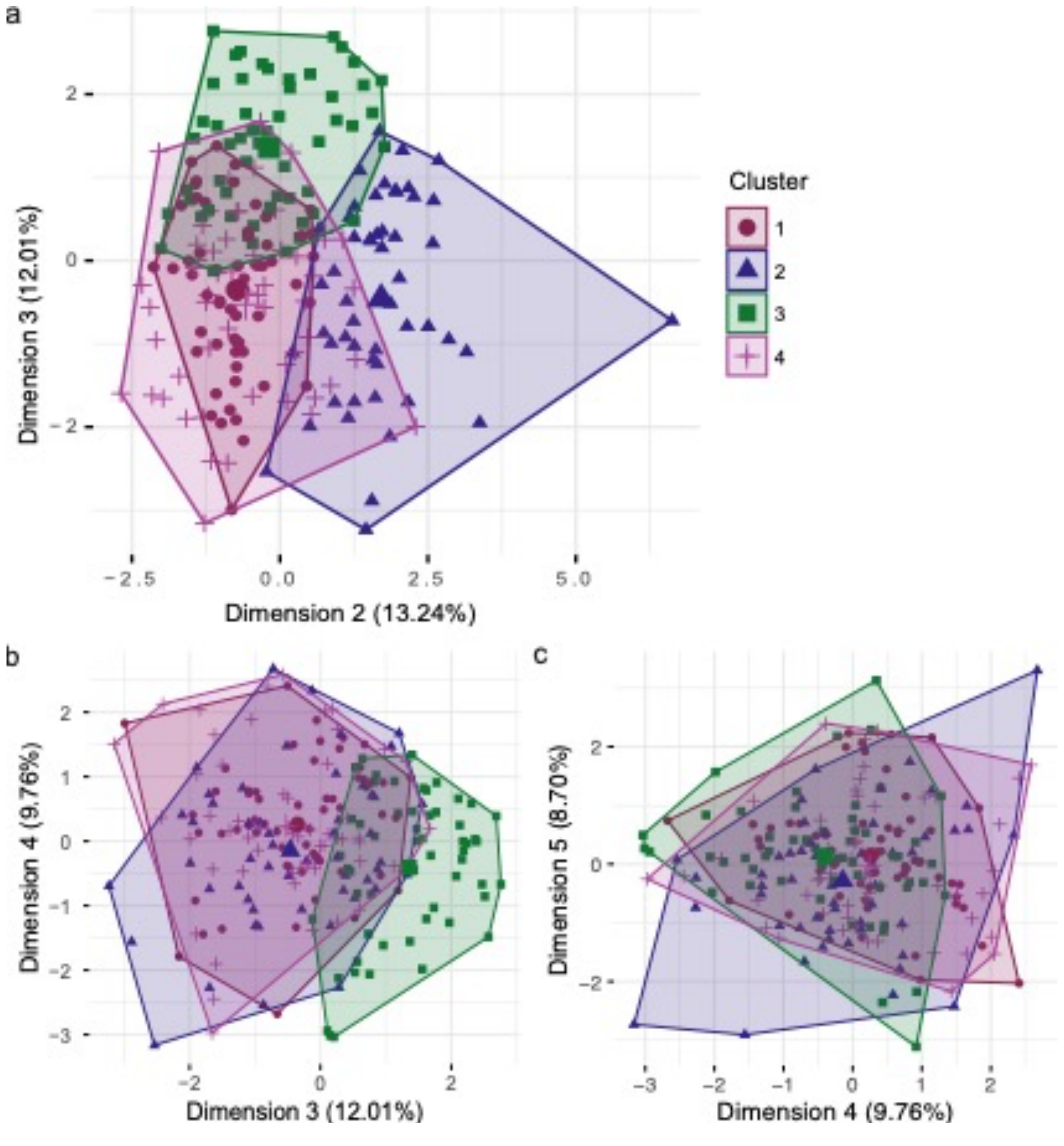

**Suppl. Figure 2** White matter lesion proportions per MS subgroup. These variables were used as input for the factor analysis. **a-h** Box plots showing the transformed proportions of white matter lesion types for 228 donors (cluster 1: n = 63; 2: n = 52; 3: n = 56; 4: n = 57). Significance was assessed by pairwise Mann-Whitney tests and adjusted for multiple testing. Only significant comparisons are shown. *p ≤ 0.05; **p < 0.01; ***p < 0.001; ****p < 0.0001. CLR = centered log ratio

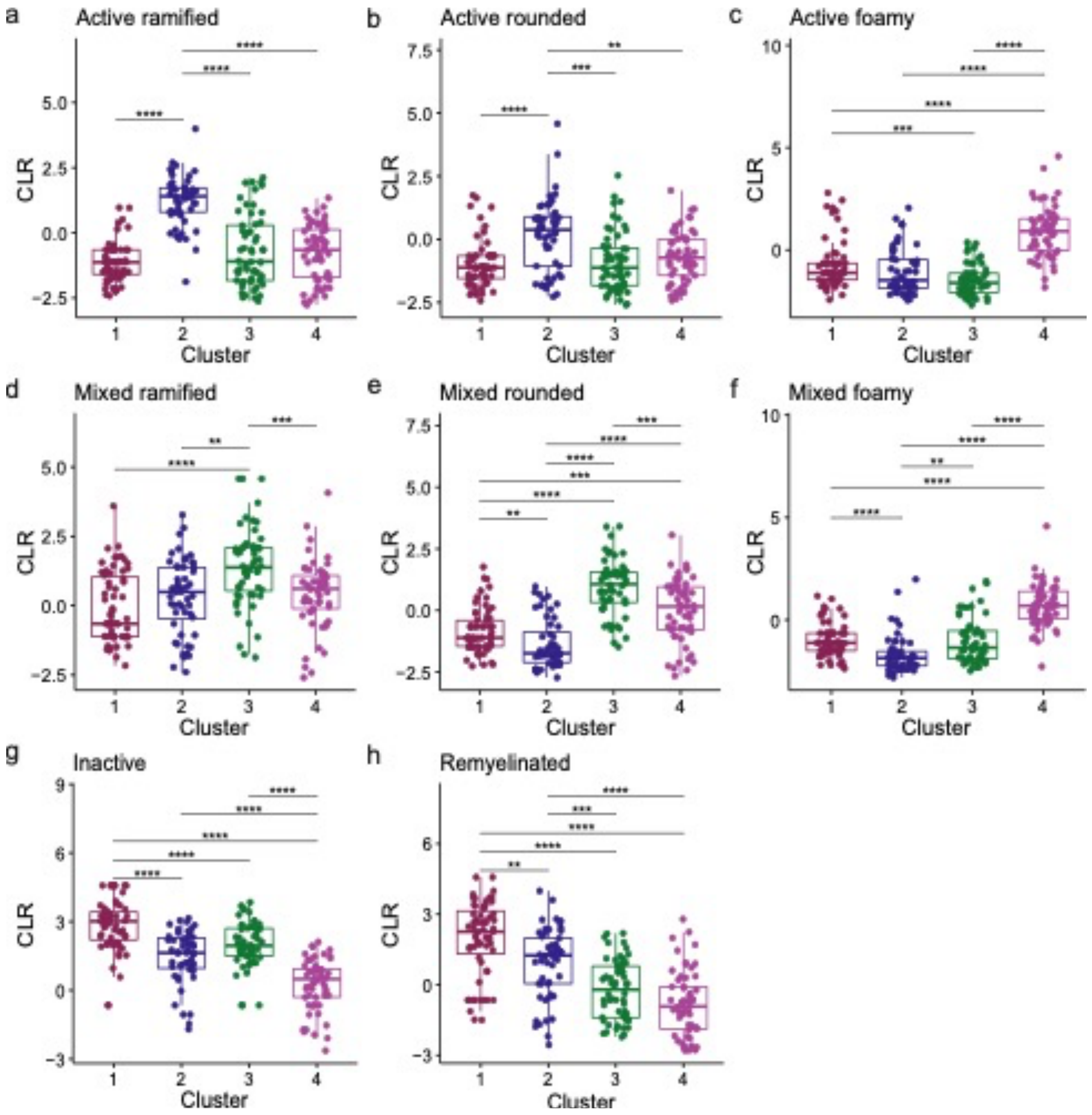

**Suppl. Figure 3** Reactive site load, lesion load and presence of nodules, cuffing and cortical lesions per MS subgroup. These variables were used as input for the factor analysis, together with the white matter lesion proportions in suppl. Figure 2. **a, b** Box plots showing the absolute number of reactive sites (**a**) and other white matter lesions (**b**) in the brainstem for 214 donors (1: n = 60; 2: n = 50; 3: n = 52; 4: n = 52). Significance was assessed by pairwise Mann-Whitney tests and adjusted for multiple testing. Only significant comparisons are shown. $^{*}p \leq 0.05$; $^{**}p < 0.01$; $^{***}p < 0.001$; $^{****}p < 0.0001$. **c** Mosaic plot showing the proportion of donors with and without nodules in any of the tissue blocks. The presence of nodules was determined for all 228 MS donors. There are significant differences between the subgroups (Fisher's Exact test; $p = 3.5 \times 10^{-4}$). Subsequent pairwise Fisher's exact tests with multiple testing correction showed that cluster 1 and 2 ($p = 6.7 \times 10^{-4}$) and 1 and 4 (p = 0.023) differ significantly. **d** Same as in **a**, but for perivascular cuffing. There are significant differences between the subgroups (p = 0.002; cluster 1 vs 4: p = 0.041 and 3 vs 4: p = 0.005). **e** Same as in **a**, but for cortical lesions. Data on the presence of cortical lesions was available for 208 donors (cluster 1: n = 58; 2: n = 51; 3: n = 48; 4: n = 51). Cortical lesions were considered absent in 32 donors (cortical tissue had been dissected without detecting lesions). Subgroups differ significantly, with donors in cluster 1 less often having (a) cortical lesion(s) ($p = 3.5 \times 10^{-8}$; cluster 1 vs 2: $p = 6.5 \times 10^{-3}$; 1 vs 3: $p = 9.8 \times 10^{-5}$; 1 vs 4: $p = 5.3 \times 10^{-6}$)

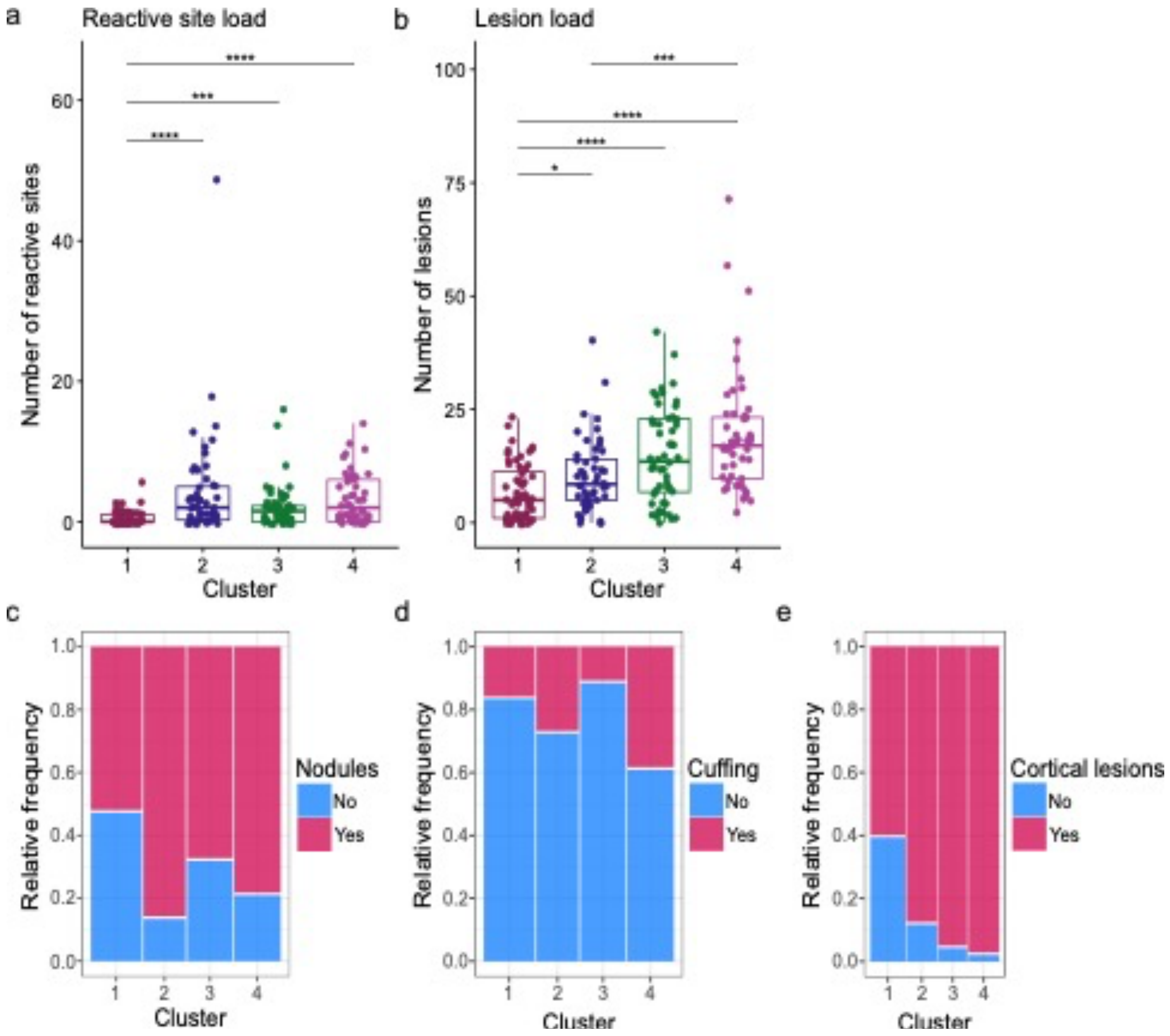

**Suppl. Figure 4** Year and cause of death (CoD) per MS subgroup. **a** Histogram of year of death. The median year of autopsy for cluster 1-4 is 2004, 2012, 2009.5 and 2008, respectively. Year of death is different between the subgroups (Kruskal-Wallis test: $p = 6.7 \times 10^{-4}$). Subsequent pairwise Mann-Whitney tests with adjustment for multiple comparisons showed that donors in cluster 2 died in later years than those in cluster 1 ($p = 1.7 \times 10^{-4}$) and cluster 4 ($p = 0.044$). **b** Bar graph of CoD for 221 donors (1: n = 63; 2: n = 48; 3: n = 54; 4: n = 56). CoD differed significantly between the subgroups (Fisher's Exact test, simulated $p \approx 0.012$), mostly with regards to the categories 'euthanasia' and '(other) natural cause'

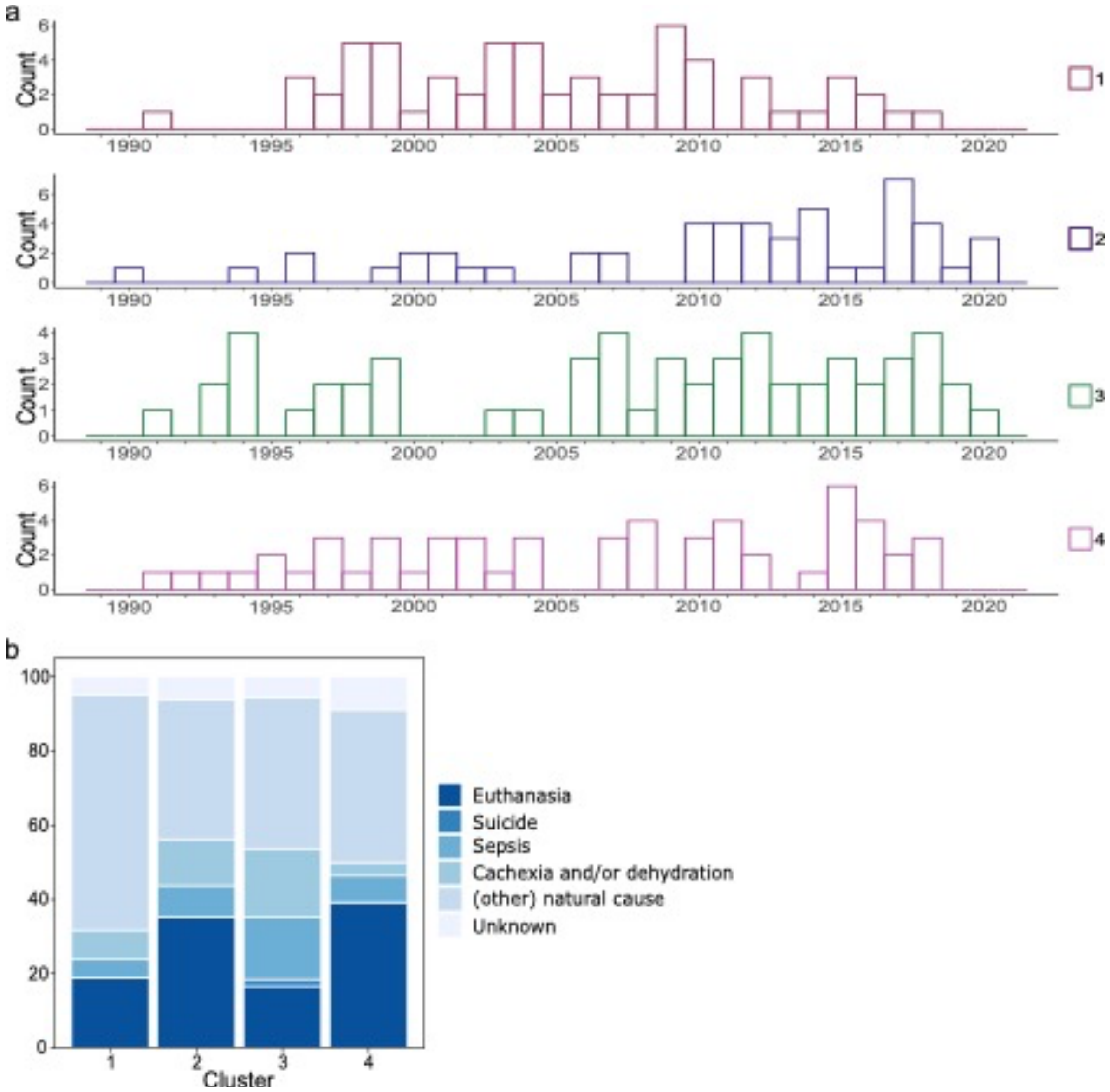

**Suppl. Figure 5** Additional neuropathological data per MS subgroup, related to the lesions in the cortex (**a**, **b**) and brainstem (**c**). **a** Box plot showing the total number of cortical lesions, for 175 donors (1: n = 35; 2: n = 45; 3: n = 46; 4: n = 49). Note that this is the same set of donors as in Figure 4a-c, with all donors having at least one cortical lesion. Significance was assessed by pairwise Mann-Whitney tests and adjusted for multiple testing. Only significant comparisons are shown. *$p \leq 0.05$; **$p < 0.01$; ***$p < 0.001$; ****$p < 0.0001$. **b** Box plot with the number of cortical lesions divided by the total number of cortical tissue blocks investigated, for 164 donors (1: n = 30; 2: n = 43; 3: n = 44; 4: n = 47). The average number of cortical lesions per tissue block does differ (Kruskal-Wallis test: p = 0.025), although subsequent pairwise Mann-Whitney tests adjusted for multiple testing did not identify significant differences between the subgroups. **c** Mosaic plot showing the proportion of donors with a lesion in the brainstem tissue block used to determine B cell presence or absence (cluster 1: n = 47 donors; 2: n = 25; 3: n = 29; 4: n = 31). There are significant differences between the subgroups (Fisher's Exact test; $p = 2.2 \times 10^{-5}$; cluster 1 vs 3: p = 0.004; 1 vs 4: $p = 6.8 \times 10^{-4}$; 2 vs 4: p = 0.017)

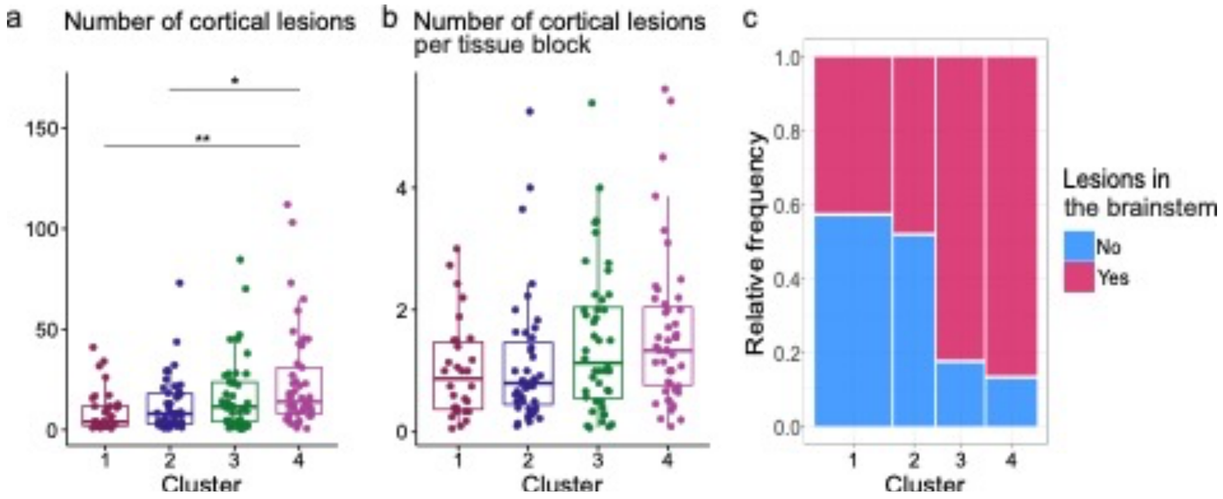

**Suppl. Figure 6** Scatter plots of MS PRS against the coordinates of donors on dimension 3 (**a**), 4 (**b**) and 5 (**c**). The fitted regression line, Pearson correlation coefficient (r) and p-value (Wald Test with t-distribution of the test statistic) are shown. The color of the dots indicates the cluster. PRS = polygenic risk score

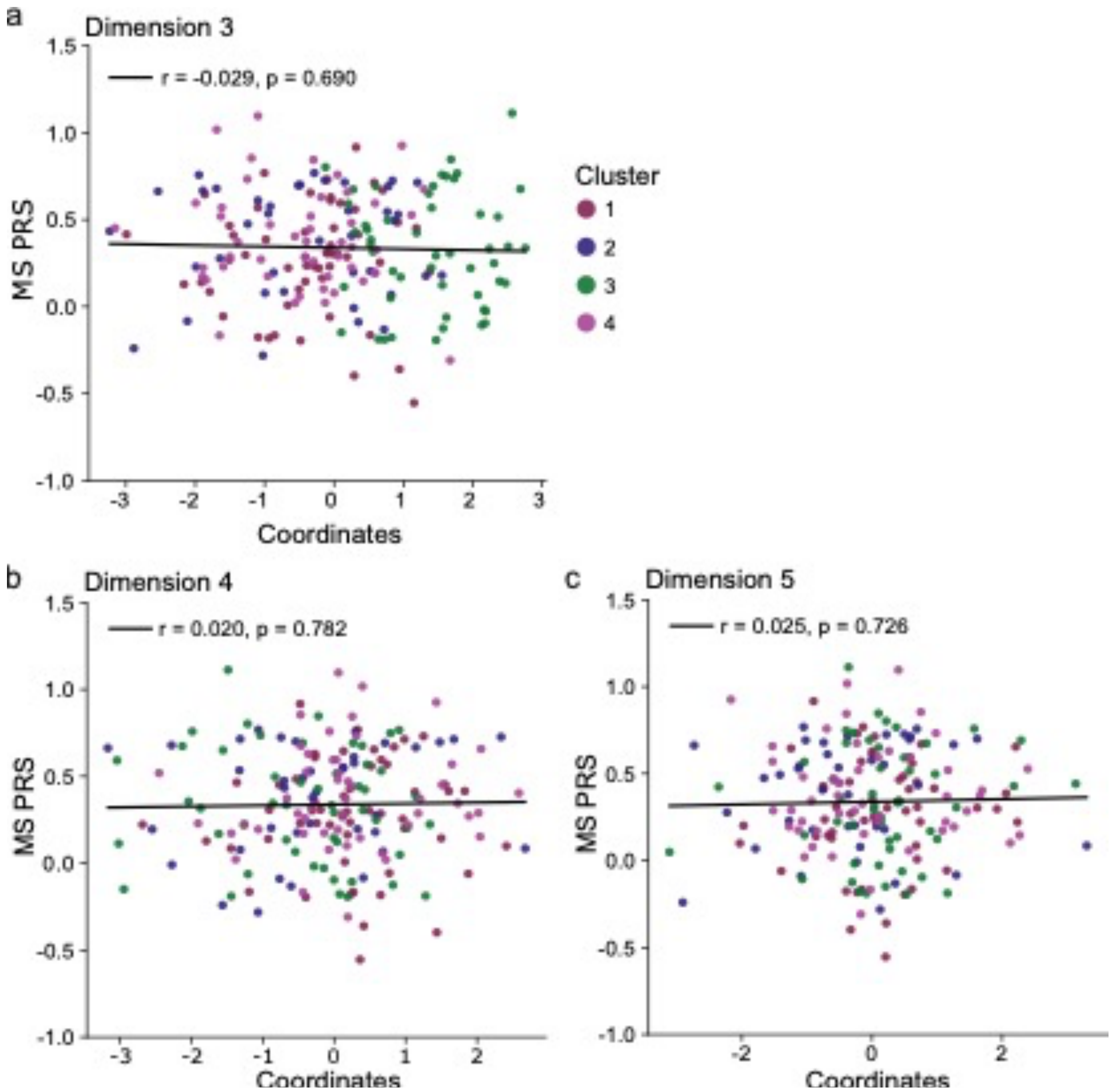
